# Cleaning and disinfecting surfaces in hospitals and long-term care facilities for reducing hospital and facility-acquired bacterial and viral infections: A systematic review

**DOI:** 10.1101/2021.12.22.21268185

**Authors:** Roger E. Thomas, Bennett C. Thomas, John Conly, Diane Lorenzetti

## Abstract

**Background:** Multiply drug-resistant organisms (MDROs) in hospitals and long-term care facilities (LTCFs) of particular concern include meticillin-resistant S*taphylococcus aureus* (MRSA), vancomycin-resistant enterococcus, multidrug-resistant *Acinetobacter* species and extended spectrum beta-lactamase producing organisms. Respiratory viruses include influenza and SARS-CoV-2.

**Aim:** To assess effectiveness of cleaning and disinfecting surfaces in hospitals and LTCFs.

**Methods:** CINAHL, Cochrane CENTRAL Register of Controlled Trials, EMBASE, Medline, and Scopus searched inception to 28 June 2021, no language restrictions, for randomized controlled trials, cleaning, disinfection, hospitals, LTCFs. Abstracts and titles were assessed and data abstracted independently by two authors.

**Findings:** Of fourteen c-RCTs in hospitals and LTCFs, interventions in ten were focused on reducing patient infections of four MDROs and/or healthcare-associated infections (HAIs). In four c-RCTs patient MDRO and/or HAI rates were significantly reduced with cleaning and disinfection strategies including bleach, quaternary ammonium detergents, ultraviolet irradiation, hydrogen peroxide vapour and copper-treated surfaces or fabrics. Of three c-RCTs focused on reducing MRSA rates, one had significant results and one on *Clostridioides difficile* had no significant results. Heterogeneity of populations, methods, outcomes and data reporting precluded meta-analysis. Overall risk of bias assessment was low but high for allocation concealment, and GRADE assessment was low risk. No study assessed biofilms.

**Conclusions:** Ten c-RCTs focused on reducing multiple MDROs and/or HAIs and four had significant reductions. Three c-RCTs reported only patient MRSA colonization rates (one significant reductions), and one focused on *Clostridioides difficile* (no significant differences). Standardised primary and secondary outcomes are required for future c-RCTs including detailed biofilm cleaning/disinfection interventions.

## Introduction

Hospitals and long-term care facilities (LTCF) are open institutions with many new admissions and discharges. The risk factors for *Clostridioides difficile* and the MDROs including meticillin-resistant S*taphylococcus* (MRSA), vancomycin-resistant enterococcus (VRE), multidrug-resistant *Acinetobacter*, resistant Gram-negative bacilli and extended spectrum beta-lactamase (R-GNB and ESBL) organisms and healthcare-acquired infections (HAIs) are exposure in a hospital or LTCF environment contaminated with these pathogens, especially a room previously occupied by an infected or carrier patient [1], age 65 and older, multimorbidity, higher severity of illnesses, and length of hospital stay [2]. Influenza and RSV have been key viral infections of concern in medical institutions for seniors and are now joined by SARS-CoV-2. The prevalence of MDROs, HAIs and *C. difficile* infection is significant in both hospitals and LTCFs.

A study of 19 acute care hospitals in Australia randomly sampled 50% (2,767) of the acute patients on 281 wards (Kappa inter-rater reliability for data entry = 0.92). The prevalence of HAIs was 9.9% (95%CI 8.8% to 11.0%), and of the 12 types of HAI found, the most frequent were surgical site 3.6% (2.9% to 4.4%), pneumonia 2.4% (1.9% to 3.1%) and urinary tract infection 2.4% (1.9% to 3.0%). The median hospital prevalence rate was 9.2% (range 5.7% to 17%). Of the 346 organisms identified in patients with an HAI, 14% were *S. aureus*, 10% *Candida albicans*, 9% *Escherichia coli* and of the 329 MDROs 113 were VRE, 101 MRSA and 67 ESB. Indwelling devices are a risk factor: in 38 patients with a bloodstream infection 35 (92%) had a vascular device in situ, of the 66 urinary tract infections 33 (50%) had an indwelling urinary catheter, and of the 41 patients with pneumonia 9 (22%) were on invasive ventilation support [4].

In a study of 28 nursing homes in southern California, bilateral samples were taken from the nares, groins and axillae of a random sample of 50 residents in each home. For 1,400 residents the median prevalence of MDRO and *C. difficile* carriage was 50% (range 24% to 70%), including MRSA 36% (range 20% to 54%); ESBL 16% (range 2% to 34%); VRE 5% (range 0% to 30%); and *C. difficile* 0% (range 0% to 8%). In addition to colonisation identified by sampling, a history of MDRO carriage was found in the medical records of 180 (13%) residents, including MRSA in 116 (8%), ESBL organisms in 81 (6%) and VRE in 22 (2%). MDRO carriage was found in 627 (45%, range 24% to 67%) patients that were previously unknown. Environmental colonisation, found in 74% of resident rooms and 93% of common areas, was assessed in each LTCF by sampling five objects in common areas (nursing station cart, table, chair, hallway handrail, and drinking fountain) and five or more objects in three resident rooms (bedside table, bedrail, call button, TV remote, door-knobs, light switch, bathroom sink and rail and flush handle) and MDROs were found in 74% of resident rooms and 93% of common areas [2].

Less is known about the survivability of human viruses on different surfaces. A systematic review of the survival of enteric viruses on soft surfaces identified 12 studies with 13 types of material and norovirus, poliovirus, rotavirus and non-human feline calcivirus and murine norovirus but the studies had no standard protocols for study size, duration of exposure, or inoculum concentration and no meta-analysis could be performed or median duration computed. Five studies of human enteroviruses concluded that (except for rotavirus) they survive better at higher ambient temperatures, and for two studies of surfaces survive longer on wool longer than on cotton with the longest survival on wool blankets at 140 days [3]. Carefully conducted studies are needed with standardised protocols and at low risk of bias testing viral survivability after disinfection interventions on materials found on patients and in rooms in hospitals and LTCFs.

A 2019 systematic review of the effectiveness of infection control programmes in LTCFs (last search 2016) identified ten c-RCTs. The interventions of four studies focused on MDROs, four on oral care to prevent respiratory tract infections, and four on hand hygiene. On the Cochrane risk of bias tool, five were considered at low risk of bias for random allocation, two for allocation concealment, eight for blinding of participants and personnel, nine for blinding of outcome concealment, nine for incomplete evidence, and ten for selective reporting. No meta-analysis was performed and the authors concluded that the infection prevention and control programmes significantly improved compliance, knowledge and quality of practices (low quality evidence) but no study assessed compliance with transmission-based precautions or implemented all five WHO core interventions [5]. The evidence for the effectiveness of cleaning and disinfection in hospitals and LTCFs requires updating in light of new developments in the rapidly evolving landscape in this area.

## Purpose

To conduct a systematic review to assess the evidence for the effectiveness of cleaning and disinfecting surfaces in hospitals and LTCFs to prevent MDROs and HAIs, including viral infections.

## Methods

The reporting of this systematic review is in accordance with PRISMA 2020 reporting guidelines for systematic reviews. The study protocol was registered with Prospero (registration number: CRD42021249823).

### Primary research question

How effective are bleach, ultraviolet light, hydrogen peroxide, copper surfaces, or copper treated fabrics at cleaning and disinfecting surfaces in hospitals and LTCFs and do they make a difference in reducing acquiring MDROs and HAIs? Our PICO statement was: **Population:** Patients in hospitals or LTCFs. **Interventions:** bleach, quaternary ammonium disinfectants, liquid or vapourised hydrogen peroxide, ultraviolet lighting, copper-plated surfaces or copper-treated fabrics, or disinfectant or antiseptic coated surfaces. **Comparison:** usual cleaning methods in the institution (or quaternary ammonium disinfectant if it is the usual method). **Outcomes:** rates of new MDROs and HAIs in patients. **Study Design:** randomized or cluster-randomized controlled trials.

### Secondary research questions

(1) What are the reported MDRO and HAI rates in the region of the institution? (2) Did the study follow or test evidence-based disinfection guidelines? (3) Did the researchers perform genomic studies to track the transmission of pathogens between the community, hospitals and LTCFs? (4) Were antibiotic stewardship programmes and their effectiveness reported? and (5) Were MDRO and HAI rates reported for environmental service workers (ESWs) and other HCWs?

### Data Collection

Five databases (CINAHL, Cochrane CENTRAL Register of Controlled Trials, EMBASE, Medline, and Scopus) were searched from inception to June 28, 2021. Searches combined keywords and database-specific subject headings from three concepts: (a) hospitals or acute/long term care facilities (e.g.: assisted living, long term care facilities, nursing homes) (b) disinfectants (e.g.: antisepsis, bleach, cleaning, copper plating of surfaces and copper impregnation of textiles, disinfection, decontamination, hydrogen peroxide, quaternary ammonium compounds, and ultraviolet (UV) light); and (c) randomized controlled trials. The Cochrane Collaboration’s highly sensitive search filter was used to identify relevant randomized controlled trials [6]. No language or date limits were applied. The reference lists of included studies were also searched to identify additional studies of relevance. The complete search strategy is documented in Appendix A.

### Study Screening

Search results were downloaded into Covidence™ [7] for de-duplication and screening. Two authors screened abstracts and full-text papers in duplicate. Disagreements were resolved through consensus. Studies were included if they were randomized or cluster controlled trials that: (1) focused on patients in hospitals or LTCFs; (2) reported on the use of bleach, quaternary ammonium disinfectants, hydrogen peroxide, ultraviolet lights, or copper-plated surfaces/copper-treated fabrics or antiseptic coated surfaces to disinfect surfaces or reduce colonisation rates in patients; and (3) reported rates of patient infection or bacterial colonisation of surfaces.

### Data Extraction and Quality Assessment

Two authors independently extracted study data and assessed the quality of all included studies. The Cochrane Risk of Bias Tool (RoB) [6] and also the Risk of Bias tool version 2 for cluster-randomized controlled trials (RoB 2) [8] were used to assess the quality of studies included in this review. All disagreements were resolved through consensus.

### Data Analysis

Two authors independently analysed study data. There were insufficient studies with enough similar study arms to permit meta-analysis.

## Results

### Search Results

The search identified 14 cluster -randomized controlled trials (c-RCTs) with clinical HAI and/or MDRO patient outcomes (ten with interventions focused on multiple bacterial pathogens, three on MRSA and one on *C. difficile*) [9-30] (Table I).

**Table I.** C-RCTs of the effects of cleaning and disinfection on infections and multiple-drug resistant infections (MDROs)

| c-RCTs of cleaning and disinfection of multiple infections |  |  |  |
| --- | --- | --- | --- |
| Author, date, setting | Randomiation, power computation, analysis | Intervention, training, detection | Outcomes |
| Anderson, 2017 [9], 2018 [10], 2018 [11], Knelson 2017 [12], Kanamori 2021 [13]<br>**Setting:** c-RCT 9 hospitals southeastern US, single rooms from which patients had been discharged who had positive cultures for any of 4 target bacteria (MRSA, VRE, *C. difficile*, or multidrug-resistant *Acinetobacter*) in previous 12 months; New admissions were 314,819 patients with 606,811 room stays of which 31,226 patients were admitted to rooms previously exposed to one of 4 target bacteria | **Randomization:** hospitals randomized to sequence of four disinfection strategies by random-number generator;<br>**Power computation:** 99% power to detect 20% decrease in incidence rates.<br>**Analysis:** GEE and ITT analysis<br>**UVC lights:** distance of UVC lights from surfaces and duration of exposure not stated<br>**Detection:** cultures of MDROs | **Intervention duration:** 28 months with four 7-month consecutive periods (1 month wash-in period then 6 months data collection).<br>**Training:** Environmental services personnel trained on use of disinfectants, cleaning protocols, UV lights; 90% compliance with protocols, UV light usage, hand hygiene, cleaning similar across study arms; all hospitals used gowns + gloves for suspected *C. difficile* or MRDO.<br>**Detection:** Standard microbiological protocols for MDRO detection, PCR nucleic acid amplification for *C. difficile* | **MDRO results:** In reference arm Quaternary ammonium (QUAD) was used (except bleach used for rooms exposed to *C. difficile* infection) 115 cases of any of 4 target organisms (= 51.3 cases/10,000 patient days of targeted organisms); UV + QUAD 33.9 cases/10,000 exposure days (RR 0.70, 95% CI 0.50–0.98; p=0.036); bleach 41.6 cases/10,000 exposure days (RR 0.85, 95% CI 0.69–1.04; p=0.116); UV + bleach 45.6 cases/10,000 exposure days (RR 0.91, 95% CI 0.76–1.09; p=0.303); *C. difficile* UV + bleach compared to bleach 30.4 cases *vs.* 31.6 cases/10,000 exposure days (RR 1.0, 95% CI 0.57–1.75; p=0.997)<br>**Hand hygiene:** QUAD study arm 89.7%; UV 88.1%, Bleach 91.4%, Bleach + UV 90.8%;<br>**Room disinfection compliance:** QUAD<br>study arm 100%; UV 95%, Bleach 100%, Bleach + UV 100%;<br>**Protocol compliance:** QUAD 100%; UV 93%, Bleach 96%, Bleach + UV 99%; UV device use 92% |
| Boyce 2017 [14]<br>**Setting:** 4 wards on 2 campuses of university-affiliated hospital in USA . Number of patients not stated, only patient bed days (10,741 in hydrogen peroxide wards, 11,490 in QUAD wards); 561 high touch surfaces sampled on peroxide wards and 575 on QUAD wards before cleaning | **Randomization:** On each campus 2 wards randomized (method not stated) to quaternary ammonia disinfectant (QUAT) and 2 to bleach. Study duration 12 months.<br>**Power computation:** None. Aerobic colony counts excluded if “fluorescent markers revealed surfaces had not been wiped or if cultures before cleaning revealed no growth.” Also “During the early months of the study, wipe rates on some study wards were as low as 52%-79%. As a result those ward months were excluded from per protocol analysis of health care outcomes.” Also “The fact that a 23% reduction in the health care-related outcomes on IHP wards was not statistically significant may have been caused in part by having to exclude a number of ward months from both the IHP and Quat arms, resulting in the per protocol analysis being underpowered to detect a statistically significant difference.”<br>**Detection:** Cultures of high touch surfaces<br>**Analysis:** mixed-effects Poisson autoregressive time-series model | **Cleaning:** Daily and discharge cleaning QUAD or bleach. In convenience sample of “several patient rooms” fluorescent marker applied to 5-8 high touch surfaces monthly. “Continued feedback” to housekeepers to maintain high wipe rates<br>**Training:** Environmental services personnel trained on use of disinfectants, cleaning protocols, UV lights; 90% compliance with protocols, UV light usage, hand hygiene, cleaning similar across study arms; all hospitals used gowns + gloves for suspected *C. difficile* or MRDO. **Detection:** Standard microbiological protocols for MDRO detection, PCR nucleic acid amplification for *C. difficile* | **Combined MRSA, VRE, and *C. difficile* rates** were assessed only during wards months when > 80% high touch surfaces had been wiped: Hydrogen peroxide cleaned wards: 86 cases/1000 patient days; QUAD cleaned wards: 119/1000 patient days; $P = 0.068$; IRR 0.77 (0.579 to 1.020)<br>**Hand hygiene compliance:** QUAD wards 95.5%, peroxide wards 95.8%<br>**Baseline:** on peroxide wards 35/561 (6.2%) of high touch surfaces had not been wiped, and 25 (4.5%) no growth before cleaning; on QUAD wards 30/575 (5.2%) 28 (4.9%).<br>**After cleaning:** Mean aerobic colony counts (ACC)/high-touch surface after cleaning peroxide (14.0 cfus), QUAD (22.2 cfus) ($P = .003$).<br>**% of surfaces no growth after cleaning:** peroxide (48%), QUAD (35%) ($P < .0001$).<br>**If select cut-off = <2.5 CFUs/cm<sup>2</sup> as definition of clean surface:** 92.4% surfaces clean after peroxide, 88.4% after QUAD ($P = .03$) |
| Chami 2012 [15]<br>**Setting:** 50 nursing homes (northwestern France; 4,345 beds, 452 new residents entered during 5 month study, 282 lost to follow-up) | **Randomization:** Homes stratified on size (> 85 residents) and Group Iso Resources score (1 = very dependent, 6 = independent), computer randomized.<br>**Power computation:** 90% power to detect 5% difference alpha 0.05 (2 tailed) required 3,524 residents.<br>**Analysis:** ITT, inflation factor 3.84 for ICC of 0.04, GEE<br>**Detection of infections:** by physician's diagnosis without requirement of laboratory confirmation | **Training:** 30 interventions (10 PPE, hand hygiene, education, sterile processes; 8 interventions for immunisation, hygiene, daily bathing, screening for incontinence, escort to bathrooms, aseptic technique; changing patient devices; 8 for preventing HAIs; 4 for identifying infections).<br>**Knowledge and compliance:** Monthly self-surveys of infection control knowledge and compliance with infection prevention guidelines | **Combined urinary, pneumonia, bronchitis, tracheobronchitis, influenza-like illness, otitis, sinusitis, rhinitis and gastrointestinal infections rate:**<br>intervention 2.11/1000 resident days, control 2.15; HR 0.99 (0.87 to 1.12, P = 0.86) |
| Heredia-Rodríguez 2018 [16]<br>**Setting:** 1,097 adult patients undergoing cardiac surgery at University Hospital, Valladolid Spain | **Intervention:** 522 patients computer randomized to receive air flow at 25 m<sup>3</sup>/hour from four ultraviolet light air sterilisers (253.7 nm), 575 to control.<br>**Power computation:** 16% infection rate in cardiac surgery, planned 10% reduction, 80% power, alpha 5%, requires 388 patients/group.<br>**Detection:** daily samples for culture from axilla, rectum, pharynx, nose, groin, chest X ray for lung infections | **Routine respiratory bundle of care to all patients:** included hand hygiene, changing of ventilator circuits when soiled or at 7 days, chlorhexidine 2% oral mouth wash every 6 hours when intubated, semi-recumbent body position at 40 degrees<br>No data on role of staff in placing and supervising air sterilisers | **Nosocomial infections:** 49 urinary tract, catheter, blood, surgical site) in intervention and 60 in control patients (ns); ventilator-associated pneumonia 4.6% vs. 5.0%, p=0.77; total infections 14.0% vs. 15.5%, p = 0.45);<br>**30-day mortality rate:** intervention 3.8%, control 6.4%) (ns) |
| Juthani-Mehta, 2015 [17]<br>**Setting:** 36 nursing homes in Connecticut; 834 residents ≥ 65 either impaired oral hygiene or difficulty swallowing | **Randomization:** Homes divided into high stratum (aides provided ≥ 140 minutes care/day) or low stratum (< 140 aide minutes) and randomized using permuted block design (method not stated) to intervention or usual care. | **Intervention:** Multicomponent pneumonia prevention intervention<br>**Staff training for intervention:** Perform for patients twice daily manual tooth and gum brushing, oral 0.12% chlorhexidine oral rinse; | **Radiographically documented pneumonia:** intervention 119 (27.4%) vs. control 94 (23.5%); HR 1.12 (.84 to 1.50, P = 0.44).<br>**Lower respiratory tract infection without radiographic documentation:** intervention 125 (28.8%), |
|  | **Power computation:** 828 participants for 25% reduction in 2.5 year pneumonia rate, alpha 0.05, power 80%, 20 % loss annually. Followed for 2.5 years; assessments by blinded study personnel;<br>**Detection:** Chest X ray for Pneumonia<br>**Analysis:** ITT, Kaplan-Meier | upright positioning during feeding | control 100 (25%); HR 1.07 (.79 to 1.46, P = .65). Data Safety and Monitoring Board terminated study for futility because conditional power nearly zero<br>**Protocol adherence:** chlorhexidine 87.9% (SD 20.5%. toothpaste 70% (SD 23.4%) |
| Lautenbach 2018 [18]<br>**Setting:** 24-bed medical ICU and a 24-bed surgical ICU (1,021 patients). No significant differences between study arms in demographics, comorbidities, indwelling devices or antibiotic use | **Randomization:** 6 beds (351 patients) in each ICU randomized (method not stated) to copper-treated linens (sheets, pillow cases and gowns), 18 beds to regular linens.<br>**Detection:** cultures for MDROs<br>**Analysis:** incidence rate ratios | **Staff training:** no statement. | **HAI rate:** 10.26/1,000 patient-days for patients with copper-treated linens and 10.41 for non-copper-treated linens, IRR = 0.99 (0.57, 1.64); P = 0.97]<br>**MDRO rate:** 3.73/1,000 patient-days for patients with copper-treated linens and 6.51 for non-copper-treated linens IRR = 0.57 (0.23, 1.26; P = 0.15]. MDRO infection and colonisation rates with use of copper linens were 43% lower but nonsignificant |
| Marik 2020 [19]<br>**Setting:** 2 ICU wards (8 beds each), 1,282 patients in eastern Virginia, USA | **Randomization:** Patients randomized (method not stated) to a ward with copper treated fabrics (sheets, pillowcases, underpads, washcloths, towels, gowns) or no copper treated fabrics.<br>**Power computation:** none;<br>**Detection:** cultures for MDROs<br>**Analysis:** rates/1000 patient days<br>**Non-randomized study in Phase 2:** both ICUs moved to a new tower, copper impregnated linens discontinued because no significant results. Workstations, tables, bathroom fittings, | Copper treated fabrics (gown, bed coverings); no statement about training of HCWs or cleaners | **Phase 1 (c-RCT):** No difference in HAI rate in intervention patients with copper impregnated linens or control patients. IRR standard vs. copper oxide linens 1.1 (0.6-2.0, p = 0.6)<br>**Phase 2: (non-randomized study):** with all patients in rooms with copper plated surfaces copper there was a significant reduction in the number of infections due to *C. difficile* (2.4 per 1000 vs. 0.7/1000 patient-days; incidence rate ratio 2.9 (95% CI 1.5 to 5.7; P = 0.0002) but no difference in the rate of central line-associated bloodstream infections or catheter- |
|  | bedrails, door handles in all rooms copper plated |  | associated urinary tract infections |
| Mitchell 2019 [20], 2018 [21], Hall 2020 [22]<br>Setting: stepped-wedge c-RCT in 11 acute care hospitals in Australia; number patients not stated; 3,534,439 occupied bed days during study | **Randomization:** by study statistician. Stepped wedge design resulted in varying intervention durations (20 to 50 weeks).<br>**Power computation;** none. **Blinding:** Hospital-wide intervention so environmental staff not blinded. Statisticians aware of timing of interventions to enable analysis. Patients “not aware.”<br>**Analysis:** Poisson generalised mixed linear models | **REACH intervention:** bundle of recommendations on optimal cleaning agents, manufacturers’ recommendations, defined cleaning frequency, training, audit, feedback, hospital-wide commitment to improve cleaning. Primary goal reduction of MRSA, *C. difficile* and VRE infections. Secondary goal thoroughness of cleaning. Nine to 16 frequent touchpoints in 50% of wards (US CDC Environmental Cleaning Checklist) in two bathrooms and bedrooms touched by invisible gel. Check of fluorescence 24 hours after cleaning | **Number of cases pre-intervention:** 230 cases VRE infection, 362 cases *S. aureus* bacteremia, 968 *C. difficile* infections for 3,534,439 occupied bed-days. During intervention 50 VRE, 109 cases *S. aureus* bacteremia, 278 *C. difficile* cases for 1,267,134 occupied bed-days.<br>**Number of cases post-intervention:** reductions in VRE infections (0.35 to 0.22/10,000 occupied bed-days; RR 0.63, 95% CI 0.41 to 0.97; p=0.0340) and non-significant reductions in *S. aureus* bacteremia cases (0.97 to 0.80/10,000 occupied bed-days; OR 0.82; 95%CI 0.60 to 1.12, p=0.2180) but not in *C. difficile* infections (2.34 to 2.52/10,000 occupied bed-days; RR 1.07; 95%CI 0.88 to 1.30, p=0.4655).<br>**Protocol compliance: % of frequent touch points cleaned:** in bathrooms increased from 55% to 76% (OR 2.07; 95%CI 1.83 to 2.34, p<0.0001) and in bedrooms from 64% to 86% (OR 1.87; 95%CI 1.68 to 2.09, p<0.0001). |
| Mody 2015 [23], Koo 2016 [24]<br>**Setting:** 418 residents with indwelling devices in 12 nursing homes, southeast Michigan | **Randomization:** NHs stratified by for profit/not for profit and computer randomized.<br>**Power computation:** 30% reduction in MDRO prevalence, power 80%, alpha .05, ICC 0.07, required 12 NHs.<br>**Detection:** cultures from nares, oropharynx, enteral feeding tube insertion site, suprapubic catheter site, groin, perianal area, and | **Training:** HCWs encouraged to perform hand hygiene before and after providing any care to participants, wear gowns and gloves when performing morning and evening care. Hand hygiene products, disposable gowns, PPE provided. 10 educational modules every 2-3 months over 3 years. Pocket cards | **MDRO prevalence density:** Intervention NHs compared to control NHs: (rate ratio, 0.77; 95%CI, 0.62-0.94); new MRSA rate ratio, 0.78; 95%CI, 0.64-0.96). for first clinically defined catheter-associated urinary tract infection HR 0.54 (95%CI, 0.30-0.97); for all (including recurrent) catheter-associated UTIs HR 0.69 (95%CI, 0.49-0.99). |
|  | wounds<br>**Analysis:** mixed-effects multilevel Poisson regression model | surveillance for infections, when to begin antibiotics.<br>**Detection:** microbiological samples all device sites at baseline, day 15, monthly up to 1 year | No reductions in new VRE or resistant gram-negative bacilli infections or in new feeding tube-associated pneumonias or skin and soft-tissue infections |
| Salgado 2013 [25]<br>**Setting:** 614 patients in ICUs in 3 hospitals in S. Carolina | **Randomization:** Bed-control service (masked to which rooms contained copper) randomly assigned (method not stated) patients to an available room, resulting in 137 patients in rooms with copper plated high touch surfaces (bedrails, overbed tables, IV poles, arms of visitor's chair; and in some nurse call button and computers) and 277 in rooms without copper plating.<br>**Power computation:** For 90% power, alpha 0.05 to detect 50% difference in health care associated infections or colonisation with methicillin-resistant MRSA or VRE required 310 patients per group. Treatment teams not masked to room assignment, HCAI evaluators masked.<br>**Detection:** cultures of high touch areas, cultures for HAIs and MDROs<br>**Analysis:** Chi<sup>2</sup>. | **Cleaning:** All rooms cleaned daily and terminal cleaning with Virex 256 (Johnson-Diversey), Dispatch (Caltech Industries) rooms with patients with *C. difficile*; Cavicide (Metrex) for spot cleaning; 46.6% of intervention patients exposed to all six copper-treated objects for entire stay, 14.3% control patients exposed to copper objects due to furniture movements.<br>**Detection:** Weekly sampling 6 sites all rooms | **HCAIs:** 89% HCAIs occurred in rooms with cumulative microbial burden > 500 colony-forming units (cfu)/100 cm<sup>2</sup>.<br>**Proportion who developed HAI and/or MRSA or VRE colonisation:** significantly lower in copper-plated rooms (0.071 vs. 0.123, p = 0.02);<br>**Proportion who developed HAI:** lower in copper-plated rooms (0.034 vs. 0.081; P = 0.013).<br>**Mortality:** copper rooms 14.3%, control rooms 15% (P = 0.64)<br>**Hand hygiene compliance:** ranged from 61% to 95%, no significant association with HAI rates (P = 0.53)<br>**Colonisation:** 4,450,545 bacteria recovered during trial; colonisation of copper surfaces ~ 6 times less likely to harbour MRSA or VRE; combined MRSA and VRE burden 96.8% lower<br>**Protocol compliance:** mean burden of standard-surface footboard in copper rooms 2,786 cfu, control 2,388 cfu (ns) |

| c-RCTs of cleaning and disinfection to reduce colonisation by a single pathogen: MRSA |  |  |  |
| --- | --- | --- | --- |
| Baldwin 2010 [26]<br>**Setting:** 793 residents ≥ 65 in 32 nursing homes in Northern Ireland | **Randomization:** NHs matched on bed numbers, staffing levels, infection control audit scores and MRSA prevalence and | **Audits:** Audits by one nurse at baseline then at 3 and 6 months and written audit reports sent to nursing home managers | **Resident MRSA colonisation prevalence:** at 12 months after intervention commenced RR 0.99 (0.69 to 1.42) |
|  | then computer randomized in pairs to intervention or control.<br>**Power computation:** for 10% reduction MRSA prevalence, assuming same colonisation rate as UK nursing homes (avg 17%), intracluster correlation 0.01, 80% power, alpha 0.05, sample size required 464 residents.<br>**Detection:** MRSA detected with PCR to detect 4 resistance genes.<br>**Analysis:** ITT, random effects meta-analysis; 40% attrition of residents | with recommendations for improvement. Swabs taken from staff nares; also resident nares, indwelling devices, urinary catheters, and wounds at baseline and at 3, 6 and 12 months. Researcher used audit tool to assess compliance with infection standards (environmental cleanliness, kitchen cleanliness, linen management, waste and sharps management, hand decontamination, PPE, urinary catheter, enteral feeding and wound management). **Training:** Staff received 2 hours of training.<br>**Training:** After each audit the intervention staff were educated and trained in hand hygiene, decontamination of equipment and environment. Selected staff in intervention NHs reinforced good infection control throughout study duration | intervention home (44/234 residents) compared to control home (47/244 residents), adjusted for clustering.<br>**Staff MRSA colonisation:** at baseline 2/169 intervention (1%), 10/164 (6%) control homes; after 12 months intervention 9/122 (7%), control 5/115 (4%)<br>**Infection control audit scores:** at baseline intervention 56% and control 53%; at 12 months (82% and 64%; $P=0.0001$). Intervention group at 12 months average audit scores attained partial (76-84%) or recommended full (> 85%) compliance in 8 out of 10 standards.<br>**Hand hygiene:** Hand hygiene (66%) and equipment decontamination (75%) still deemed poor in the intervention group. Main problems consistently identified were excess jewelry, false and long nails, and lack of hand decontamination after removal of gloves and aprons. |
| Bellini 2015 [27]<br>**Setting:** 104 nursing homes (4750 residents) in Canton Vaud, Switzerland, | **Randomization:** NHs randomized by random-number generator to decolonisation intervention or control (standard precautions).<br>**Power computation:** 90% if assumed 20% MRSA prevalence in control NHs, 70% decolonisation success rate; 51% if assumed 20% success rate and 20% increase in controls. Achieved power was 51%.<br>Universal baseline and 12 | **Staff training:** All staff control and intervention NHs sessions on standard infection precautions. Patient Intervention: topical decolonisation nostrils, pharynx, skin, hair, dental prostheses, MRSA ulcers x 5 days. Decolonisation assessed as successful if two MRSA swabs negative 7 days apart; if unsuccessful then decolonisation was repeated.<br>Environmental | **NH screening rates:** intervention 87% (range 20-100%), control 86% (range 27-100%). Average 87% residents accepted screening.<br>**MRSA prevalence at baseline:** intervention 8.9% (range 0-38%), control 8.9% (range 0-43%).<br>**MRSA prevalence of colonisation with intervention:** Intervention rate 8.9% declined to 5.8% ($P=0.003$) after intervention, control 8.9% |
|  | month screening residents for MRSA;<br>**Detection:** cultures from nostrils, groin, ulcers and urine(if permanent catheter)<br>**Detection:** HAIs assessed by cultures<br>**Analysis:** GEE | decontamination: clothes, linen, bed, bedside table, phone, walking aid, wheelchair armrests, and TV control | to 6.6% ($P = .02$); no difference between arms. |
| Peterson 2016 [28], Schora 2014 [29]<br>**Setting:** 12 nursing units in 3 long term care facilities, Illinois (862 beds, 4,200 annual admissions, 365,809 patient days) | **Randomization:** random assignment of units by statistician;<br>**Power computation:** >80% power for 50% reduction in disease prevalence in intervention arm (2-sided $\alpha = .05$), ICC $\leq 0.03$<br>**Detection:** Cultures of nares and wounds<br>**Analysis:** $\chi^2$ and Fisher exact test for proportions, Poisson tests for rates | **(1) Universal screening for MRSA at beginning of study** then 5 cycles (a) MRSA screening, with (b) universal decolonisation with intranasal mupirocin and a chlorhexidine bath twice (1 month apart) and (c) environmental decontamination all flat surfaces in rooms, common rooms and equipment with bleach over a 1 week period at start of intervention period.<br>**(2) All new admissions screened** with real-time PCR (polymerase chain reaction), and MRSA positive patients decolonised but not isolated.<br>**(3) Enhanced cleaning flat surfaces** with bleach wipe every 4 months.<br>**(4) Training:** Annual pathogen transmission, hand hygiene, disinfection education instruction. | 16,773 swab tests performed over 2 years.<br>**MRSA infection rate baseline year 1:** 44/365,809 resident days;<br>**Unit colonisation rate:** baseline 16.64%. During study year 1 intervention units 11.61% ($P = .028$), control units 17.85% ($P = .613$), difference ($P = .001$).<br>**MRSA infection rate year 2:** 12/287,847 resident days (decrease compared to baseline $p < .001$); During study year 2 RCT was discontinued “due to patient commingling,” all units received intervention, colonisation rate 10.55% (difference between baseline and year 2 ($P < .001$); decrease 65% ($P < .001$) and in each LTCF ($P < .03$). Most conversions occurred on short stay units |

| **c-RCT of cleaning and disinfection to reduce colonisation by a single pathogen: *C. difficile*** |  |  |  |
| --- | --- | --- | --- |
| Ray 2017 [31]<br>**Setting:** 16 acute care hospitals northeastern Ohio, USA. Number of residents not stated.<br>Power computation based on 1,068 cases and 1,683,928 patient days in | **Randomization:** Hospitals stratified into 4 groups on *C. difficile* rates then randomized (method not stated) to intervention or usual cleaning;<br>**Power computation:** 95% power to detect 25% reduction *C difficile*; | **Cleaning:** All hospitals used bleach wipes to daily disinfect *C. difficile* rooms.<br>**Training:** intervention arm environmental services personnel training on cleaning, daily individual feedback on | **Protocol compliance:** Intervention hospitals fluorescent marker removal post-discharge both *C. difficile* and non-*C. difficile* rooms: increased over 12 months 63% (hospitals ranged 23% to 74%) to 82% (range 64% to |
| facilities in 2009 | **Detection:** all hospitals used nucleic acid tests for diagnosis; samples from bed rail, table, call button, telephone for 3 months before study commenced and then for 12 months of study; cultures for *C. difficile*<br>**Analysis:** linear mixed model | cultures and removal of fluorescent markers (5 surfaces for daily cleaning, 10 surfaces for post-discharge cleaning in each hospital for 10 *C. difficile* rooms and 10 rooms not *C. difficile*) | 100%).<br>**For daily cleaning of *C. difficile* rooms marker removal:** increased 52% (hospitals range 12% to 74%) to 69% (range 35% to 91%), $P < .001$.<br>***C. difficile* cultures:** For intervention hospitals CDI rooms positive cultures for *C. difficile* declined 13% to 3% ($P < .01$).<br>*C. difficile* rooms in control hospitals: no significant reductions in positive *C. difficile* cultures (hospitals ranged 19% to 14%, $P = .23$).<br>**Incidence of *C. difficile*/10,000 patient days comparing intervention and control hospital arms:** No significant differences in intervention or post-intervention periods<br>**Percentage of positive cultures after cleaning *C. difficile* rooms and non-*C. difficile* rooms and HCAI;** no correlation |

### 14 C-RCTs: Numbers of patients studied

Of the 14 c-RCTs, eleven reported the number of patients (total 329,254 patients), Mitchell [20] reported 3,434,439 occupied bed days but not patient numbers, Boyce also reported on 22,231 occupied bed days but not patients [14] and Ray reported basing the power computation on 1,068 cases and 1,683,928 patient days in the study facilities in 2009 but reported neither patients nor occupied bed days for the actual study period [30]. The largest studies by far were Anderson (31,226 patients exposed to MDROs and a total of 329,254 in all hospitals studied) [9-13] and Mitchell (3,534,439 bed-days) [20].

### 10 C-RCTs with patient outcomes and studied multiple infections

Ten of the c-RCTs studied varying interventions with outcomes of acquisition of MDROs or HAIs and three (Anderson [9-13], Mitchell [20-22], and Salgado [25]) found decreased HAIs or MDROs or both after cleaning of surfaces as a major component and one (Mody [23, 24]), with enhanced education. The largest c-RCT (Anderson [9-13] studied 31,226 patients in nine hospitals in the southeastern US who were admitted to single rooms which had been terminally disinfected after the discharge of a previous occupant with positive cultures for any of four target microorganisms (MRSA, VRE, *C. difficile*, or multidrug-resistant *Acinetobacter*) in the previous 12 months. The 11 hospitals had a total of 314,819 patients and these patients were also assessed for wider circulation of MDROs. The new admissions were randomized to rooms which had received one of four disinfection strategies. After completion of the interventions to reduce the environmental presence of the target MDROs and *C. difficile*, a primary composite outcome of new hospital-acquired colonisation or infection with these microorganisms was studied. The lowest rates of MDRO/*C. difficile* acquisition for the new admissions were after using ultra-violet light (UV) + quaternary ammonia detergent (QUAD) with 33.9 cases/10,000 exposure days which was statistically significant result (RR 0.70, 95% CI 0.50–0.98; p=0·036) compared to the standard cleaning arm. Cleaning with bleach revealed 41.6 cases/10,000 exposure days, with UV + bleach 45.6 cases/10,000 exposure days and then for the reference study arm (QUAD) 51.3 cases/10,000 patient days. For *C. difficile* infection alone, for bleach and for UV + bleach, there were 30.4 cases and 31.6/10,000 exposure days, respectively. Staff hand-hygiene compliance averaged 90%, room disinfection compliance 100%, and protocol compliance was QUAD 100%; and in study arms UV arm 93%, Bleach arm 96%, Bleach + UV arm 99%; and UV device use arm 92% [QUAD is also referred to as QUAT in some publications] (Table I).

The next largest c-RCT (Mitchell) was conducted in 11 acute care hospitals in Australia. The number of patients involved was not stated but the study involved 3,534,439 occupied bed days. The focus was on increasing cleaning thoroughness with the REACH bundle: using optimal cleaning agents, following manufacturers’ recommendations, defined cleaning frequency, training, audit, feedback, and a hospital-wide commitment to improve cleaning. There was a significant reduction in the primary outcomes of VRE clinical infections from 0.35/1,000 occupied bed days to 0.22 but no significant decrease in MRSA or *C. difficile* infections. There were significant but insufficient increases in the secondary outcome of frequent touch points cleaned in bathrooms from 55% to 76% and in bedrooms from 64% to 86% [20-22].

Salgado randomized 614 patients in ICUs in three hospitals in South Carolina either to rooms with copper surfaces on high touch surfaces (bedrails, overbed tables, IV poles, arms of the visitor’s chair, and in some the nurse call button and computers) or to rooms without copper plating. Forty-six patients (7.5%) developed HAIs and 26 (4.2%) became colonised with MRSA or VRE. For the composite primary outcome of HAI or colonisation with MRSA or VRE the rate was significantly lower in copper-plated rooms (0.071 vs. 0.123, p = 0.02). For HAI only, the incident rate was reduced from 0.081 to 0.034 (P = .013). Copper surfaces were ∼ 6 times less likely to harbour MRSA or VRE and the combined MRSA and VRE burden was 96.8% lower. Hand hygiene compliance ranged from 61% to 95% and was not significantly associated with HAIs [25].

Mody in a three-year study randomized 418 residents in 12 nursing homes in southeast Michigan who had indwelling devices either to the intervention group in which HCWs were encouraged to perform hand hygiene before and after providing any care to participants and wear gowns and gloves when performing morning and evening care, or to a usual care control group. No specific new cleaning agents or disinfection protocols were deployed in this study but rather enhanced education and precautions. Training consisted of 10 educational modules every 2-3 months over 3 years. There were no significant reductions in the intervention group for the primary outcome related to overall prevalence density of MDROs: MRSA, VRE or resistant gram-negative bacilli organisms. The secondary outcome of any device associated infection revealed significant reductions in first catheter-associated urinary tract infection occurrence (RR 0.54, 95%CI 0.30 to 0.97) and all catheter-associated infections (RR 0.69, 95%CI 0.49 to 0.99) but not for new feeding tube–associated pneumonias or skin and soft-tissue infections [23,24].

Six c-RCTs which aimed to reduce multiple MDRO acquisitions or HAI infections did not find significant reductions in their respectively designated primary outcomes. Boyce in a study of four wards on two campuses of a university-affiliated hospital in Connecticut did not report the number of patients but a total of 22,231 patient days. The wards were randomized each month during the 12-month c-RCT either to use liquid hydrogen peroxide or continue usual daily and terminal cleaning with quaternary ammonium disinfectant (QUAD). There were non-significant decreases in the primary composite outcome of patients with MRSA and VRE colonisation or infection and *C. difficile* infections. After peroxide cleaning the combined colonisation of room surfaces by MRSA, VRE, or hospital onset *C. difficile* was lower with average aerobic colony counts (ACC)/high-touch surface of 14.0 cfus and after QUAD 22.2 cfus (*P* = .003). The percentage of surfaces with no growth after cleaning was higher for peroxide (48%) than QUAD (35%; *P* < .0001). The authors proposed a cut-off of <2.5 CFUs/cm^2^ as the definition of a clean surface, and 92.4% of surfaces were clean after peroxide and 88.4% after QUAD (*P* = .03) with this criterion. [14].

Chami in a cluster randomized study of 4,345 residents in 50 nursing homes in France used 30 interventions (PPE, hand hygiene, education, sterile processes, immunisation, hygiene, daily bathing, screening for incontinence, escort to bathrooms, aseptic technique, changing patient devices, preventing HAIs, and identifying infections) and for the primary outcome of the combined urinary, upper and lower respiratory and gastrointestinal infection rate found no significant differences between the intervention and control arms [15]. No specific new cleaning agents or disinfection protocols were deployed in this study.

Heredia-Rodriguez in a study at the university hospital, Valladolid, Spain, provided all cardiac surgery patients with a routine respiratory bundle of care including hand hygiene, changing ventilator circuits when soiled or at seven days, chlorhexidine 2% oral mouth wash every six hours when intubated, and semi-recumbent body position at 40 degrees, then randomized 522 to air flows exposed to UV light sterilisers and 575 to no UV light air conditioning and found no significant differences in ventilator-associated pneumonia or all infections [16].

Juthani-Mehta in 36 nursing homes in Connecticut with 834 residents randomized 434 patients to a multicomponent pneumonia prevention programme (twice daily manual tooth and gum brushing and 0.12% chlorhexidine oral rinse and upright positioning during feeding) and 400 to usual care. Adherence to the chlorhexidine protocol was 87.9% (SD 20.5%) and 100% for upright feeding. There were no significant differences in pneumonia rates over the 2.5 years of the intervention [17]. No specific new cleaning agents or disinfection protocols were deployed in this study

Lautenbach in a study of 1,021 patients in two 24-bed ICUs in Philadelphia randomized patients to use copper-treated linens (fitted sheets, sheets, pillow cover and gown) or usual linens but found no significant differences in MDRO rates. The authors commented that the study was likely underpowered [18]. Marik in a study of 1,282 patients in two ICU wards (8 beds in each) in eastern Virginia randomized the intervention group to use copper-treated linens (sheets, pillowcases, underpads, wash cloths, towels and gowns) or usual linens and there was no difference in HAIs [19].

### Four C -RCTs studied single infections

Four c-RCTs focused on single pathogens. Ray studied HAI *C. difficile* infection in a study of 16 acute care hospitals in northeastern Ohio and reported both CDI outcomes and surface colonisation rates. He did not state the number of residents but the power computation was based on 1,068 cases and 1,683, 928 patient-days in the facilities in 2009. All hospitals used bleach wipes to daily disinfect *C. difficile* rooms. In the intervention arm environmental services workers (ESWs) received training on cleaning and daily individual feedback on cultures and how many fluorescent markers were removed after cleaning. There was an average increase in fluorescent marker removal over 12 months from 63% (hospitals ranged from 12% to 74%) to 69% (range 35% to 91%) and a significant decrease in *C. difficile* colonised rooms positive for *C. difficile* from 13% to 3% in the intervention group and no significant changes in the control group but there were no significant differences in the intervention or post-intervention periods in the incidence of *C. difficile*/10,000 patient days [30].

Three c-RCTs reported only MRSA colonisation rates. Peterson in three LTCFs in Illinois with 4,200 annual admissions randomized 12 units either to perform universal MRSA decolonisation with intranasal mupirocin and a chlorhexidine bath twice and decontamination of all flat surfaces in rooms, common rooms and equipment with bleach and enhanced cleaning of flat surfaces with bleach wipe every four months, or to usual care. New admissions were screened and decolonised. All nursing staff were trained in pathogen transmission, hand hygiene, and disinfection education. The baseline colonisation rate was 16.84% and after one year 11.61% in the intervention units (p = 0.028) and after one year 17.85% in the control units (p = 0.61) [28,29].

Baldwin in a study of 793 residents in 32 nursing homes in northern Ireland randomized 16 units to a staff infection control and education programme to reduce MRSA colonisation and 16 to usual practice. Audits were conducted at baseline, 3 and 6 months and written audit reports were sent to nursing home managers with recommendations for improvement. There was 40% attrition of residents during the study. In the intervention group over 12 months infection audit control scores increased from 56% to 84% and in the control group from 53% to 64%, but hand hygiene (66%) and equipment decontamination (75%) were still poor in the intervention group and in an ITT analysis there were no significant differences in MRSA colonisation rates between the intervention and control group after 12 months [26].

Bellini in a study of 4,750 residents in 104 nursing homes in Switzerland randomized 53 homes to the intervention group and 51 to the usual care control group. There were wide baseline ranges in MRSA rates between nursing homes: in the intervention group the nursing home average was 8.9% (range 0-38%) and in the control group the nursing home average was 8.9% (range 0-43%). In the intervention group all HCWs participated in training sessions on the concepts and practice of standard precautions that should be applied to all residents independent of their MRSA status. In the intervention group all patients received topical decolonisation of their nostrils, pharynx, skin, hair, dental prostheses and MRSA ulcers and patients’ clothes, linen, bed, bedside table, phone, walking aid, wheelchair armrests, and TV control were decolonised. The MRSA rate in the intervention arm declined from 8.9% to 5.8% (P =.003) and also in the control from 8.9% to 6.6% (P = .02) but there were no significant differences between study arms [27].

### Secondary outcomes

Of the 14 included c-RCTs one reported an increase in MRSA rates in their region (eastern Switzerland) at the time of the study [27] (personal communication from Dr. Bellini). Baldwin identified a recently reported UK nursing home study with an MRSA prevalence of 17% and based his power computation on that report but did not report trends in the institutions he studied or the region [26]. Anderson did not report trends in their hospital region but identified that the addition of UV light to standard chemical disinfection in the rooms previously occupied by MDRO patients led to a significant reduction in risk for the overall group of 11 hospitals of acquiring *C. difficile* (RR 0.89; 95%CI 0.80 to 0.99; p = 0.031) and VRE (RR 0.56 (0.31 to 0.996; p = 0.048) [14,15]. Mitchell commented that “First, Australia has major reservoirs of *C. difficile* outside the hospital environment. Secondly, genetically diverse strains of *C. difficile* are being transmitted into hospitals and infecting patients.” [20].

Four c-RCTs referenced guidelines. Mitchell assessed their pragmatic research design against the PRagmatic-Explanatory Continuum Indicator tool (described in detail in the article [22] and appendices) and based the high touch points sampled on the CDC Environmental Cleaning Checklist. Chami in her study of 25 NHs in France presented during HCW training a”Delphi web survey of guidelines.” [15]. Mody in her study of 17 NHs in southeast Michigan reported that all Medicare-certified and Medicaid-certified NHs have an infection control program and a part-time preventionist and her Targeted Infection Prevention Intervention (TIP) included barrier precautions, surveillance and feedback for NH staff education as key infection prevention principles [23,24]. Salgado referenced the CDC Guideline for Disinfection and Sterilization in Healthcare Facilities 2008 but the study intervention was copper surfaces, a new intervention [25]. No guidelines were referenced by the other studies (Table I).

One study reported that in each of the three LTCFs the MRSA clone (pulsotype) USA !00 predominated (range 38% to 92.3%) and there were no clonal associations between the mupirocin-resistant MRSA strains [28]. Mitchell did not perform microbiological testing of the environment or whole genome sequencing because of financial constraints [20]. Anderson stated “we did not do molecular analyses to confirm that organisms included in our outcomes were related to organisms in the environment as this task was impossible given the scope of our study.” [9].

Mitchell reported that all hospitals had an antibiotic stewardship programme throughout the study period but did not report its results and found during the pre-intervention period a combined antibiotic medication monthly defined daily dose of 4,614/1000 occupied bed days and during the intervention period was 3,949/1000 [20]. Baldwin reported that the consumption of antibiotics in the previous 12 months was 12% in the control and 14% in the intervention group [26] and Bellini in the previous 30 days was 11% ± 5.6 in the control and 14% ± 8.1 in the intervention group but neither provided antibiotic stewardship data [27].

Only one C-RCT reported MDRO rates in HCWs. Baldwin reported that MRSA rates in cleaners were 6% in the intervention and 1% in the control group but did not report results for ESWs [26].

#### Meta-Analysis

There was significant heterogeneity amongst the studies which precluded meta-analysis.

### Risk of bias

Using the Cochrane Risk of Bias Tool, of the 12 c-RCTs which reported patient MDROs and or/HAIs, six were at low risk of bias for randomization (all used computer programmes) and six were unknown as they merely stated they were randomized. For allocation concealment two were at low, nine unknown and one at high risk. However, the cleaning and disinfection interventions were applied to room surfaces and not to patients by the environmental service workers employed by the hospitals and LTCFs. The thoroughness of disinfection was assessed by environmental service supervisors (ESS) employed by the hospitals and LTCFs and also by the research staff and feedback was provided by the ESSs to the environmental service workers. For blinding of participants and personnel four were at unknown and eight at high risk. For blinding of outcome assessors four were at low, four unknown and four at high risk. However, for both blinding assessments, considering that interventions were applied to all surfaces in predefined clusters using hospital and LTCF patient registries, and outcomes are laboratory-confirmed infections with MDROs or HAIs the risk of bias due to blinding was revised to low. For attrition eight were at low and four at unknown risk. Although several very large studies had a constant throughput of patients during the study period, the tracking of patients by patient bed occupancy which used the hospital admission and discharge system is likely to find all participants. All 12 studies were at low risk of selective reporting (Figures 1 and 2). The overall assessment is low risk but high for allocation concealment.

**Figure 1.**
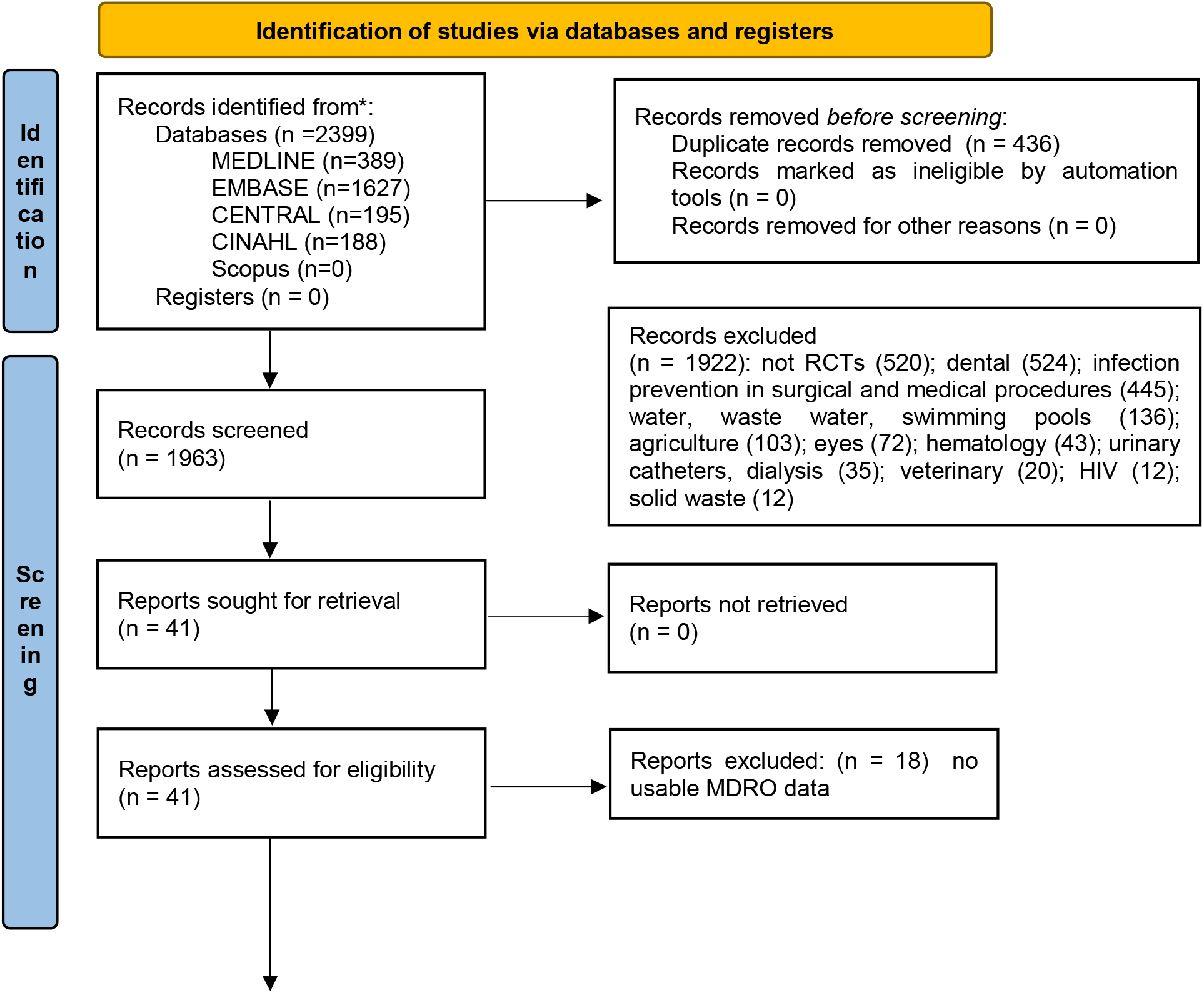
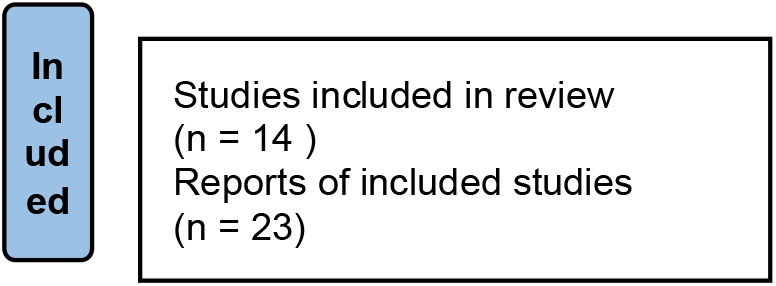
PRISMA Flow diagram for new systematic reviews which include searches of databases and registers only.

**Figure 2.**
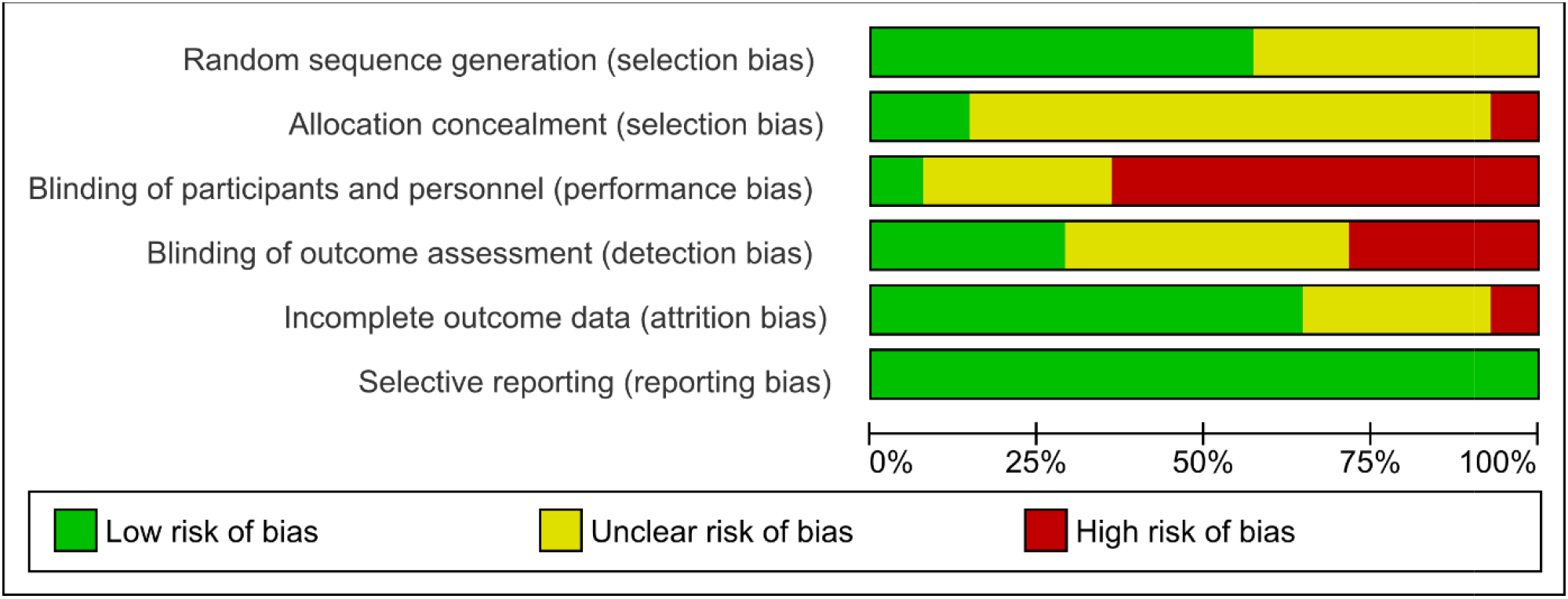
Cochrane Risk of Bias Graph

**Figure 3.**
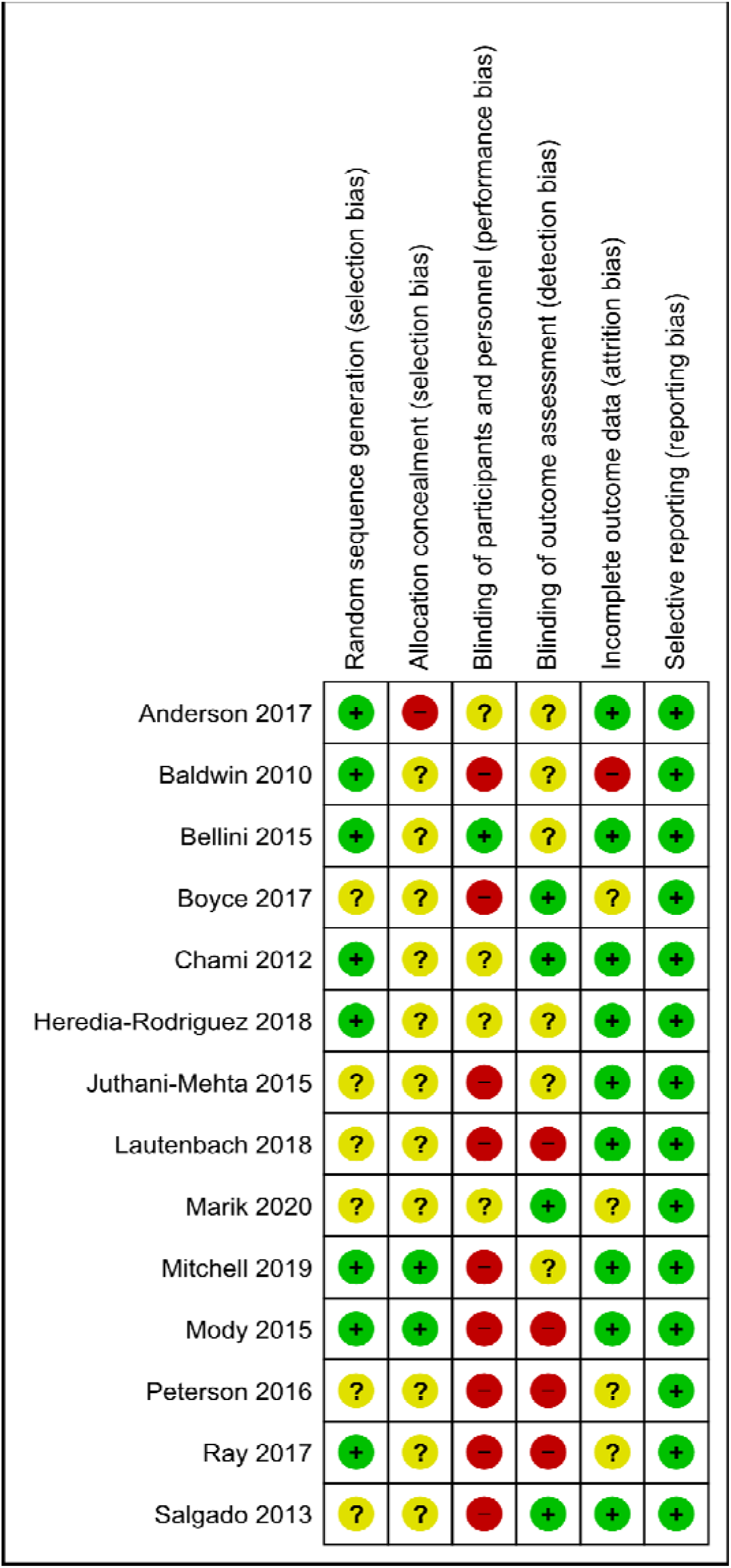
Cochrane Risk of Bias Summary

Considering separately the four c-RCTs which reported patient colonisation rates Boyce [14] and Ray [30] also reported patient outcomes (reported above). For randomization three were at low and one at unknown risk, for allocation concealment four were at unknown risk, for blinding of participants and personnel one was at low risk and three at high risk, for blinding of outcome assessors one was at low, two unknown and one at high risk, and for incomplete data one was at low, two unknown and one at high risk, and for selective reporting all four were at low risk. The same comments that the interventions were provided to predetermined clusters of wards by nursing staff and/or environmental service workers employed by the hospitals and LTCFs apply also to these colonisation studies and thus the overall assessment is revised to low risk but risk was high for allocation concealment. Four studies required ongoing patient cooperation for personal decolonisation measures [15,16, 27,28]. Twelve c-RCTs reported a power computation. (Appendix B).

Six c-RCTs reported corrections for clustering. Key questions in the Cochrane RoB version 2 tool for c-RCTs are: 1. Are there baseline differences between clusters? Ten c-RCTs provided detailed tables comparing the intervention and control groups and showed no significant baseline differences; 2. Were all participants identified and recruited before randomization? In all 14 c-RCTs these processes occurred before randomization; 3. Were there any deviations from Protocols? In 10 c-RCTs the interventions were applied directly to the patients’ environment automatically, and in four the interventions were applied to the patients and followed Protocols with no reported exceptions, and 4. Were people delivering the intervention aware of the participant’s assigned intervention during the trial? Because the interventions usually lasted from one to three years it is likely that in the four where interventions were applied directly to patients by HCWs this knowledge was likely apparent to HCWs and could have been apparent to patients (Appendix B). Nevertheless, the GRADE assessment was low risk (Appendix C).

## Discussion

Only five of the 14 c-RCTs reviewed identified a reduction in their *a priori* identified primary outcomes. Of these five studies, three looked at cleaning and disinfecting agents including UV, copper and bleach as outlined in our primary research question and the other two looked at mutlimodal educational tools. Anderson’s study compared four cleaning methods and found the lowest MDRO rates in the ultraviolet light + quaternary ammonia detergent (QUAD) arm (33.9 MDRO cases/10,000 exposure days) with the next lowest rate in the bleach arm (41.6 cases/10,000 exposure days) and the highest in the QUAT reference arm (51.3 cases/10,000 exposure days). However the rate in the UV + bleach arm was higher than in the UV light + QUAD at 45.6 cases/10,000 exposure days [9-13]. Salgado cleaned surfaces with three disinfectants then randomized patients to rooms with copper-plated surfaces (bedrails, overbed table, IV poles, and arms of the visitor’s chair). For a composite primary outcome of new HAI or VRE/MRSA colonisation Salgado found significant differences: 21 cases/294 patients (7%) in rooms with copper-plated surfaces and 41/320 (13%, p = 0.02) in rooms without copper-plated surfaces [25}. For HAI only, the rate was reduced from 0.081 to 0.034 (P = .013). Peterson used decontamination strategies with both patients (mupirocin to nares and open wounds and a chlorhexidine body wash) and the environment (decontamination of surfaces in all patient and common rooms and equipment with bleach). The average baseline MRSA colonisation rate was 16.64% and there was a significant reduction in the intervention (11.61%, P = .028) but not in the control units (17.85%, P =.613) difference between intervention and control P =.001 [28].

The other two studies focused on educational and organisational strategies which demonstrated reductions in their primary outcomes although Mitchell’s study also measured the completeness of implementation of the REACH bundle of optimal cleaning agents, following manufacturers’ recommendations for their use, defining cleaning frequency, training, audit, feedback, and an explicit hospital-wide commitment to improve cleaning. Mitchell designed the implementation framework using the Promoting Action on Research Implementation in Health Services (i-PARIHS) framework study and measured implementation with the Standards for Reporting Implementation Studies (StaRI) framework [27]. In the intervention arm the number of cases of VRE/10,000 occupied bed days (OBD) declined from 0.65 to 0.43 (RR 0.63 (95%CI 0.41 to 0.97, p= 0.034) and although rates of *C. difficile* declined from 2.74/OBD to 2.19, *S. aureus* bacteraemia from 1.02 to 0.86, MSSA from 0.84 to 0.69, and MRSA from 0.19 to 0.17 none were significant reductions [20-22]. Mody found significant reductions in four outcomes compared to the control for all MDROs (RR 0.77; 95%CI, 0.62-0.94, p <.05); new MRSAs (RR 0.78; 95%CI, 0.64-0.96, p <.01), first clinically defined catheter-associated urinary tract infection (HR 0.54 (95%CI, 0.30-0.97, p <.04); and all catheter-associated UTIs (including recurrent) (HR 0.69 (95%CI, 0.49-0.99, p = 0.045) but no reductions in new VRE, R-GNB, new feeding tube–associated pneumonias or skin or soft-tissue infections [23].

The other nine c-RCTs found no significant results but the majority of these studies with the exception of Chami [15] and Bellini [27] had higher risk of bias. Of these studies Boyce [14], Heredia-Rodríguez [16], Juthani-Mehta [17], Lautenbach [18], Marik [19], and Ray [30] reported on the clinical outcomes of HAI in either a composite outcome or as a stand-alone outcome. Only two of these studies reported on the use of novel cleaning and disinfecting agents and the majority focused on training and education of healthcare workers.

No institution reported completely eradicating MDROs or HAIs. How close to zero the number of cfus remaining after disinfection should be has not been defined and only one of the included studies proposed a definition of a clean surface at <2.5 cfu/cm^2^ [14]. The relationship between the density of cfus per specified surface area and the risk of infection is likely to vary with the performance of the patient’s immune system, the amount and duration of exposure, and with the frailness and multimorbidity of the patients. The studies did not provide explanations why there were often substantial differences in MDRO baseline rates between institutions in their studies.

Hospitals and LTCFs are open to admissions of urgent and non-urgent patients and how referrals between LTCFs and hospitals with a region contribute to MDROs and HAIs has neither been systematically explored nor has the contribution of symptomatic and asymptomatic patients, staff and visitors. Key solutions will be to genomically measure MDROs and HAIs and track their transmission patterns, and comprehensively and rapidly screen all admissions and use isolation strategies until patients are cleared, which will require an increase in isolation units and the staff to screen patients. ESWs are crucial workers, their training and evaluation need to be improved, turnover rates minimised and pay increased to reduce MDRO and HAI rates. There is inadequate research on the degree to which contamination of cleaning cloths and solutions and continued use on multiple surfaces transfer contamination to other surfaces.

### Circulation of pathogens between hospitals, nursing homes and the community

None of the 14 c-RCTs assessed the circulation of pathogens as patients transferred between hospitals, LTCFs and the community. If future studies perform these assessments asymptomatic carriers could be identified to permit better strategies to isolate patients who bring MDROs into institutions. A four-year study of whole genome sequencing of 1,250 symptomatic *C. difficile* cases in Oxfordshire, UK, found that 45% of the cases were genetically different from previous cases and a possible environmental source of contamination could be identified for only 2% of patients [31,32]. Bellini’s study of Swiss nursing homes found that the MRSA incidence correlated with LTCF size and inversely with staff/resident ratios and thus LTCF size, multiple patient occupancy of rooms and transfer rates to and from hospitals may affect the predominant clones and affect the difficulty of the decolonisation tasks [27].

### Antibiotic stewardship

Mitchell reported that all hospitals had an antibiotic stewardship programme but did not describe the protocol or its operation for any of the hospitals. Mitchell’s was the only study which reported pre-intervention and intervention antibiotic use and for all 11 acute care hospitals. During the pre-intervention period the combined monthly defined daily dose was 4,614/1000 occupied bed days and during the intervention period was 3,949/1000. There was a wide range both pre-intervention at the 25% prescribing level (786/1000) and the 75% level (6,182/1000) and also post-intervention at the 25% level (2,090/1000) and the 75% level (5,957/1000) [20]. Antibiotic stewardship programmes and thorough supervision of their effectiveness are important in reducing MDRO colonisation. High rates of antimicrobial use were reported in Russo’s large study of 19 Australian hospitals in which 44% of patients were receiving antibiotics (excluding surgical prophylaxis) and 10% were being treated for at least one MDRO [3].

### Environmental cleaning staff

Only one study reported staff MRSA colonisation rates although it is known that staff hands are often colonised [27]. The ESW staff are key to attaining thoroughness in disinfection of surfaces and several authors commented on the high turnover of ESW staff. Only Mitchells’ REACH study stated the numbers of ESWs (1,729) [21] but no c-RCT stated the number of rooms and surfaces cleaned per shift so it is unknown if the ESWs’ workload makes thorough cleaning per protocol feasible. Studies varied in the amount and frequency of feedback to cleaners. No study identified differences in effectiveness between individual cleaners, the effect of educational interventions or factors associated with superior cleaning outcomes, or understanding of the language used for training, educational materials and feedback. There are no studies of merit pay for superior cleaning outcomes. Some studies cleaned all rooms, some only known colonised rooms, the number of high touch points cleaned varied between studies, and these issues are likely to affect the overall effectiveness of ESW staff. The difficulty of cleaning LTCF rooms filled with personal possessions has not been addressed. Deterioration of surfaces making cleaning more difficult was commented on by one study [34]. The cleaning methods, diligence, duration, amount of pressure used, and type of cleaning cloths and disinfectants used will affect outcomes and need detailed comparative studies. Studies are needed to compare for individual ESWs the differences in effectiveness and the causes of heterogeneity among ESWs in relationship to their training, supervision and difficulty of the environments they are asked to disinfect. There are no economic evaluations to assess how the intensity and effectiveness of cleaning reduce total medical costs.

### Mask-wearing and hand hygiene

Mask wearing and hand hygiene are key interventions to prevent person-to-person transmission of viral respiratory infections and the most comprehensive and recent systematic review is the 2020 Cochrane review by Jefferson [34] and it will need updating for studies during the COVID-19 pandemic period. In many studies the risk of bias for the RCTs and c-RCTs was high or unclear. The review included only three c-RCTs of hand hygiene in LTCFs. McConeghy’s 2017 study was assessed at unclear risk for random sequence generation and allocation concealment, and high risk for blinding of personnel and participants, outcome assessment and selective reporting [35]. Temime’s 2018 study was found to be at high risk for random sequence generation, unclear risk from allocation concealment, high risk from blinding of participants and personnel, performance assessment and incomplete data [36] while Yeung’s 2011 study was considered at unclear risk from random sequence generation, allocation concealment, selective reporting and at high risk regarding blinding of participants and personnel and outcome assessment [37]. There was only one c-RCT of hand hygiene for patients and staff which assessed bacterial contamination and there was a significant decrease in bacterial colonisation in the intervention group.

A 2020 systematic review of hand washing, distancing and mask wearing to prevent transmission of SARS-CoV-2 identified 172 observational studies in 16 countries but no RCTs and a meta-analysis of 44 non-randomized studies in health-care and non-health-care-settings (25,697 individuals) found that the odds ratio of virus transmission at distances more than one metre was 0.18 (0.09 to 0.38, moderate certainty) compared to distances < 1 metre, for mask wearing OR was 0.15 (0.07 to 0.34), but with low certainty of evidence [38].

#### Research on which surfaces are most frequently touched by patients, staff and visitors and which surface textures are most colonised by biofilms

The included trials included a variable number of surfaces which they designated as high touch: bed rails, overbed tables, bedside tables, nurse call bells, visitor chairs, keyboards, bathrooms, toilets, and toilet floors. Mattresses and mattress covers were not sampled. Medical equipment cleaning was usually considered a nursing responsibility. The frequency, duration and firmness of contact were not assessed. Surface types (plastic, stainless steel, copper) and textures (shiny, rough, composite) were not assessed for differential rates of bacterial or biofilm contamination. The frequency, duration and firmness of cleaning contact, techniques employed (horizontal rubbing, widening circles) and types of cleaning cloths and mops (composition, number of uses on individual surfaces before disposal) used by ESWs were not assessed. The presence of biofilm formation on any types of surfaces was not assessed. All of the areas identified above related to the types of surfaces, cleaning techniques and their effect on bacterial contamination and biofilms are research gaps which need to be addressed.

#### Trials in economies in transmission and developing economies

No trials of the work of ESWs were identified in economies designated by the United Nations as economies in transition or developing economies. Thus it is not known what the workload of ESWs is in these hospitals, which surfaces they clean, the nature of those surfaces, how supervision and measurement of the effectiveness of disinfection are undertaken, and MDRO and HAI rates in relationship to ESW activities. We were unable to find any publications in our searches that addressed the surveillance of MDROs and HAIs in relationship to ESW work and the adequacy of laboratory facilities to measure bacterial contamination and ESW disinfection effectiveness.

## Conclusions and Future Directions

Although there were some signals suggesting the use of novel strategies as adjuncts to cleaning and disinfection including UV and copper the results of examining the c-RCTs to date remain heterogenous and no definitive findings were found related to UV or copper with some studies associated with a reduction in outcomes and others not, depending on the setting. We found that “composite” outcomes seemed to be used often and only certain marker microorganisms were used. There were also major differences between primary and secondary outcomes. It would be useful to develop a set of standardised primary and secondary outcomes for future studies to enable comparisons between studies. A recent systematic review on the use of copper impregnation of clothing and commonly touched surfaces suggested there was no significant advantage to the use of this strategy [39]. The use of UV light in conjunction with usual cleaning agents and disinfectants deserves further study.

The key requirements to reduce MRDO and HAI rates are comprehensive screening of patients, genomic studies to trace infection transmission when available, rapid assessment and quarantining of admissions with additional isolation rooms and assessment staff, improved cleaning methods (especially biofilms because none of the included c-RCTs assessed biofilms), and comprehensive approaches to cleaning and disinfecting surfaces and objects in hospitals and LTCFs which can harbour and transmit microorganisms.

## Strengths

The literature search had no restrictions of language or date. The risk of bias was assessed with both the Cochrane risk of bias tool and the Cochrane risk of bias tool version 2 for cluster-RCTs. The total number of patients was large (11 studies with 329,254 patients) and for the three studies which reported only patient-occupied bed days 5,140,598 bed-days. Anderson’s 31,226 patients are included in the total above and he also assessed infection rates for another 329,254 patients in the nine study hospitals not directly targeted by the interventions [11]. The outcome was laboratory-confirmed MDROs or HAIs for 13/14 studies and clinically confirmed infections in one study [15].

## Weaknesses

Eight studies were at low risk of bias for randomization and the other six only mentioned that they were “randomized.” Only two studies confirmed that they concealed randomization. Blinding of personnel, patients and data assessors was low but the rating was graded low risk because patients were randomized in pre-defined clusters and the outcome was laboratory-confirmed infections. It is recognised there are significant difficulties in blinding patients to the use of various cleaning agents.

Minimal information was available for the five secondary outcomes: (1) reporting of regional MDRO and HAI infections before the study commenced, (2) following or testing an evidence-based guideline to reduce infections, (3) performing genomic studies to identify the transmission of infections between the community, hospitals and LTCFs, (4) reporting antibiotic use and antibiotic stewardship before and during the intervention period, and (5) measuring the MDRO and HAI rates in ESW and other healthcare staff.

## Supporting information

PRISMA Statement

## Data Availability

All data produced in the present study are available upon reasonable request to the authors

## Author Contributions

**C**onceptualisation RET and BCT, literature searches RET and DLL, data entry, risk of bias analyses and data analysis RET and CBT, text RET, text editing and revision RET, JC and DLL.

## Institutional Review Board Statement

Systematic review so not needed.

## Informed Consent Statement

Systematic review so not needed.

### Appendix A. Literature Search

#### MEDLINE(R) and Epub Ahead of Print, In-Process, In-Data-Review & Other Non-Indexed Citations and Daily <1946 to June 28, 2021> (OVID)

1. exp Nursing Homes/ or Long-Term Care/ or Homes for the Aged/ or Assisted Living Facilities/ or exp Hospitals/ or Adolescent, hospitalized/ or Child, hospitalized/ or Inpatients/
2. exp Hospital Units/ or Patients’ Rooms/ or exp Equipment/ and Supplies, Hospital/ or exp Surgical Equipment/
3. (assisted living or care home* or convalescence facilit* or convalescence home* or elder* care facilit* or extended care or (home adj3 aged) or (long term adj2 care) or nursing home* or old age facillit* or old age home* or old folks home* or (residential adj2 care) or retirement facilit* or retirement home*).tw,kf.
4. (hospital* or inpatient* or (patient* adj2 room*) or unit or units or ward or wards).tw,kf.
5. 1 or 2 or 3 or 4
6. exp Antisepsis/ or Infection control/ or Sterilization/ or Disinfection/ or Disinfectants/ or Decontamination/ or Housekeeping/
7. (antisepsis or asepsis or aseptic or clean* or decontaminat* or disinfect* or housekeep* or sanitis* or sanitiz*).tw,kf.
8. 6 or 7
9. Ultraviolet Rays/
10. (UV or ultra-violet or ultraviolet).tw,kf.
11. Sodium Hypochlorite/
12. Chlorine/
13. Hydrogen Peroxide/
14. Bleaching Agents/
15. exp Quaternary Ammonium Compounds/
16. Copper/
17. (albone or antiformin or bleach* or chlorine or clorox or copper* or crystacide or dihydrogen dioxide or hydrogen dioxide or hydrogen peroxide or hydrogenperoxide or hydroperoxide or quaternary ammoni* or quaternary bisammoni* or quaternized amine or sodium hypochlorite).tw,kf.
18. 9 or 10 or 11 or 12 or 13 or 14 or 15 or 16 or 17
19. 5 and 8 and 18
20. (randomized controlled trial or controlled clinical trial).pt.
21. (groups or placebo or randomized or randomly or trial).tw,kf.
22. 20 or 21
23. 19 and 22
24. animals/ not humans/
25. 23 not 24

#### Embase <1974 to 2021 June 28> (OVID)

1. nursing home/ or long term care/ or home for the aged/ or exp hospital/ or exp hospital patient/
2. exp “laboratory and hospital equipment”/ or exp medical device/
3. (assisted living or care home* or convalescence facilit* or convalescence home* or elder* care facilit* or extended care or (home adj3 aged) or (long term adj2 care) or nursing home* or old age facillit* or old age home* or old folks home* or (residential adj2 care) or retirement facilit* or retirement home*).tw,kw.
4. (hospital* or inpatient* or (patient* adj2 room*) or unit or units or ward or wards).tw,kw.
5. 1 or 2 or 3 or 4
6. asepsis/ or antisepsis/ or infection control/ or instrument sterilization/ or exp disinfection/ or disinfection system/ or disinfectant agent/ or decontamination/ or cleaning/ or housekeeping/
7. (antisepsis or asepsis or aseptic or clean* or decontaminat* or disinfect* or housekeep* or sanitis* or sanitiz*).tw,kw.
8. 6 or 7
9. exp ultraviolet radiation/
10. (UV or ultra-violet or ultraviolet).tw,kw.
11. hypochlorite sodium/
12. chlorine/
13. hydrogen peroxide/
14. exp bleaching agent/
15. exp quaternary ammonium derivative/
16. copper/
17. (albone or antiformin or bleach* or chlorine or clorox or copper* or crystacide or dihydrogen dioxide or hydrogen dioxide or hydrogen peroxide or hydrogenperoxide or hydroperoxide or quaternary ammoni* or quaternary bisammoni* or quaternized amine or sodium hypochlorite).tw,kw.
18. 9 or 10 or 11 or 12 or 13 or 14 or 15 or 16 or 17
19. 5 and 8 and 18
20. randomized controlled trial/ or controlled clinical trial/ or randomization/ or intermethod comparison/
21. (placebo* or random*).tw,kw.
22. (compare or compared or comparison).ti.
23. ((evaluated or evaluate or evaluating or assessed or assess) and (compare or compared or comparing or comparison)).ab.
24. (open adj label).tw.
25. ((double or single or doubly or singly) adj (blind or blinded or blindly)).tw.
26. double blind procedure/
27. parallel group$1.tw.
28. (crossover or cross over).tw.
29. ((assign$ or match or matched or allocation) adj5 (alternate or group$1 or intervention$1 or patient$1 or subject$1 or participant$1)).tw.
30. (assigned or allocated).tw.
31. (controlled adj7 (study or design or trial)).tw.
32. (volunteer or volunteers).tw.
33. human experiment/
34. trial.ti.
35. 20 or 21 or 22 or 23 or 24 or 25 or 26 or 27 or 28 or 29 or 30 or 31 or 32 or 33 or 34
36. 19 and 35
37. (rat or rats or mouse or mice or swine or porcine or murine or sheep or lambs or pigs or piglets or rabbit or rabbits or cat or cats or dog or dogs or cattle or bovine or monkey or monkeys or trout or marmoset$1).ti. and animal experiment/
38. Animal experiment/ not (human experiment/ or human/)
39. 37 or 38
40. 36 not 39

#### EBM Reviews - Cochrane Central Register of Controlled Trials <May 2021>

1. exp Nursing Homes/ or Long-Term Care/ or Homes for the Aged/ or Assisted Living Facilities/ or exp Hospitals/ or Adolescent, hospitalized/ or Child, hospitalized/ or Inpatients/
2. exp Hospital Units/ or Patients’ Rooms/ or exp Equipment/ and Supplies, Hospital/ or exp Surgical Equipment/
3. (assisted living or care home* or convalescence facilit* or convalescence home* or elder* care facilit* or extended care or (home adj3 aged) or (long term adj2 care) or nursing home* or old age facillit* or old age home* or old folks home* or (residential adj2 care) or retirement facilit* or retirement home*).tw
4. (hospital* or inpatient* or (patient* adj2 room*) or unit or units or ward or wards).tw
5. 1 or 2 or 3 or 4
6. exp Antisepsis/ or Infection control/ or Sterilization/ or Disinfection/ or Disinfectants/ or Decontamination/ or Housekeeping/
7. (antisepsis or asepsis or aseptic or clean* or decontaminat* or disinfect* or housekeep* or sanitis* or sanitiz*).tw
8. 6 or 7
9. Ultraviolet Rays/
10. (UV or ultra-violet or ultraviolet).tw
11. Sodium Hypochlorite/
12. Chlorine/
13. Hydrogen Peroxide/
14. Bleaching Agents/
15. exp Quaternary Ammonium Compounds/
16. Copper/
17. (albone or antiformin or bleach* or chlorine or clorox or copper* or crystacide or dihydrogen dioxide or hydrogen dioxide or hydrogen peroxide or hydrogenperoxide or hydroperoxide or quaternary ammoni* or quaternary bisammoni* or quaternized amine or sodium hypochlorite).tw
18. 9 or 10 or 11 or 12 or 13 or 14 or 15 or 16 or 17
19. 5 and 8 and 18
20. animals/ not humans/
21. 19 not 20

***************************

### CINAHL (EBSCO 1982 to June 28 2021)

1. ((MH “Nursing Homes+”) OR (MH “Nursing Home Patients”) OR (MH “Long Term Care”) OR (MH “Assisted Living”) OR (MH “Housing for the Elderly”) OR (MH “Hospitals+”) OR (MH “Hospital Units+”) OR (MH “Patients’ Rooms+”) OR (MH “Emergency Service+”) OR (MH “Inpatients”) OR (MH “Aged, Hospitalized”) OR (MH “Adolescent, Hospitalized”) OR (MH “Infant, Hospitalized”) OR (MH “Child, Hospitalized”) OR (MH “Ancillary Services, Hospital”) OR (MH “Residential Facilities”)) OR TI ((assisted living or care home* or convalescence facilit* or convalescence home* or elder* care facilit* or extended care or (home N3 aged) or (long term N2 care) or nursing home* or old age facillit* or old age home* or old folks home* or (residential N2 care) or retirement facilit* or or retirement home*)) OR AB ((assisted living or care home* or convalescence facilit* or convalescence home* or elder* care facilit* or extended care or (home N3 aged) or (long term N2 care) or nursing home* or old age facillit* or old age home* or old folks home* or (residential N2 care) or retirement facilit* or or retirement home*)) OR TI ((hospital* or inpatient* or unit or units or ward or wards)) OR AB ((hospital* or inpatient* or unit or units or ward or wards))
2. ((MH “Sterilization and Disinfection+”) OR (MH “Disinfectants”) OR (MH “Asepsis”) OR (MH “Housekeeping Department”) OR (MH “Cleaning Compounds”)) OR TI ((antisepsis or asepsis or aseptic or clean* or decontaminat* or disinfect* or housekeep* or sanitis* or sanitiz*)) OR AB ((asepsis or aseptic or clean* or decontaminat* or disinfect* or housekeep* or sanitis* or sanitiz*))
3. ((MH “Ultraviolet Rays”) OR (MH “Sodium Hypochlorite”) OR (MH “Chlorine”) OR (MH “Hydrogen Peroxide”) OR (MH “Quaternary Ammonium Compounds+”) OR (MH “Copper”)) OR TI ((albone or antiformin or bleach* or chlorine or clorox or copper* or crystacide or dihydrogen dioxide or hydrogen dioxide or hydrogen peroxide or hydrogenperoxide or hydroperoxide or quaternary ammoni* or quaternary bisammoni* or quaternized amine or sodium hypochlorite)) OR AB ((albone or antiformin or bleach* or chlorine or clorox or copper* or crystacide or dihydrogen dioxide or hydrogen dioxide or hydrogen peroxide or hydrogenperoxide or hydroperoxide or quaternary ammoni* or quaternary bisammoni* or quaternized amine or sodium hypochlorite)
4. 1 and 2 and 3
5. (MH “Randomized Controlled Trials”) OR (MH “Single-Blind Studies”) OR (MH “Triple-Blind Studies”) OR (MH “Double-Blind Studies”) OR (MH “Random Assignment”) OR (MH “Pretest-Posttest Design+”) OR (MH “Cluster Sample”) OR (MH “Placebos”) OR (MH “Crossover Design”) OR (MH “Comparative Studies”)
6. (MH “Sample Size”) AND AB ((assigned OR allocated OR control))
7. TI ((randomised OR randomized or trial)) OR AB (random*)
8. AB (control W5 group) OR AB (cluster W3 RCT)
9. 5 or 6 or 7 or 8
10. 4 and 9
11. Limit 4 to Publication Type: Randomized Controlled Trial
12. 10 or 11

#### SCOPUS

(TITLE-ABS-

KEY ((antisepsis OR asepsis OR aseptic OR clean* OR decontaminat* OR disinfect* OR housekeep* OR sanitis* OR sanitiz*)) AND TITLE-ABS-

KEY ((albone OR antiformin OR bleach* OR chlorine OR clorox OR copper* OR crystacide OR dihydro gen AND dioxide OR hydrogen AND dioxide OR hydrogen AND peroxide OR hydrogenperoxide OR hydroperoxide OR quaternary AND ammoni* OR quaternary AND bisammoni* OR quaternized AND amine OR sodium AND hypochlorite)))

### Appendix B. Cochrane Risk of Bias tool for cluster-randomised trials (ROB – 2)

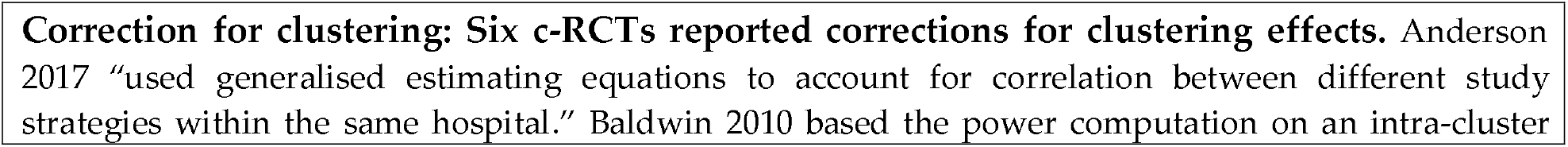
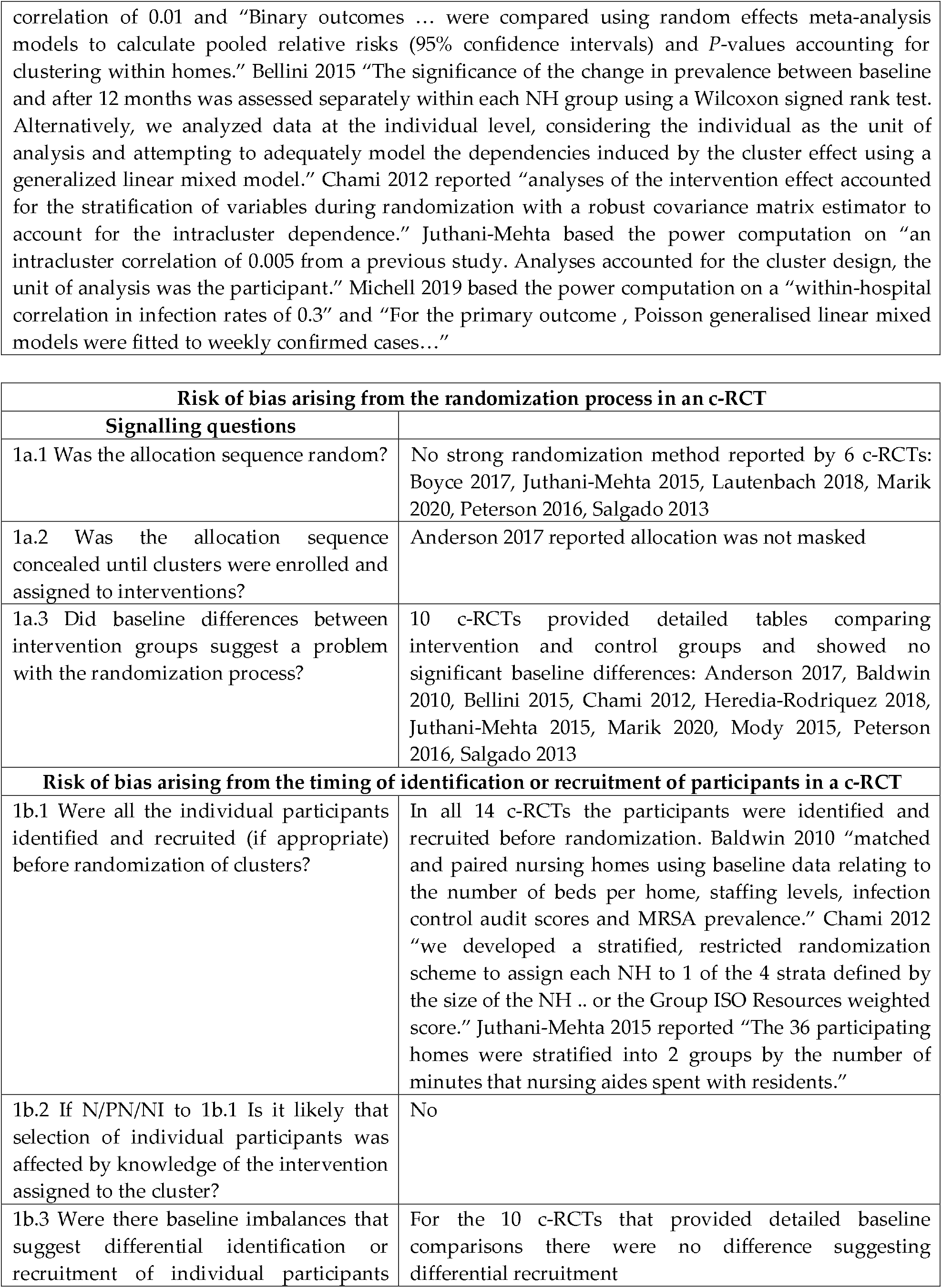
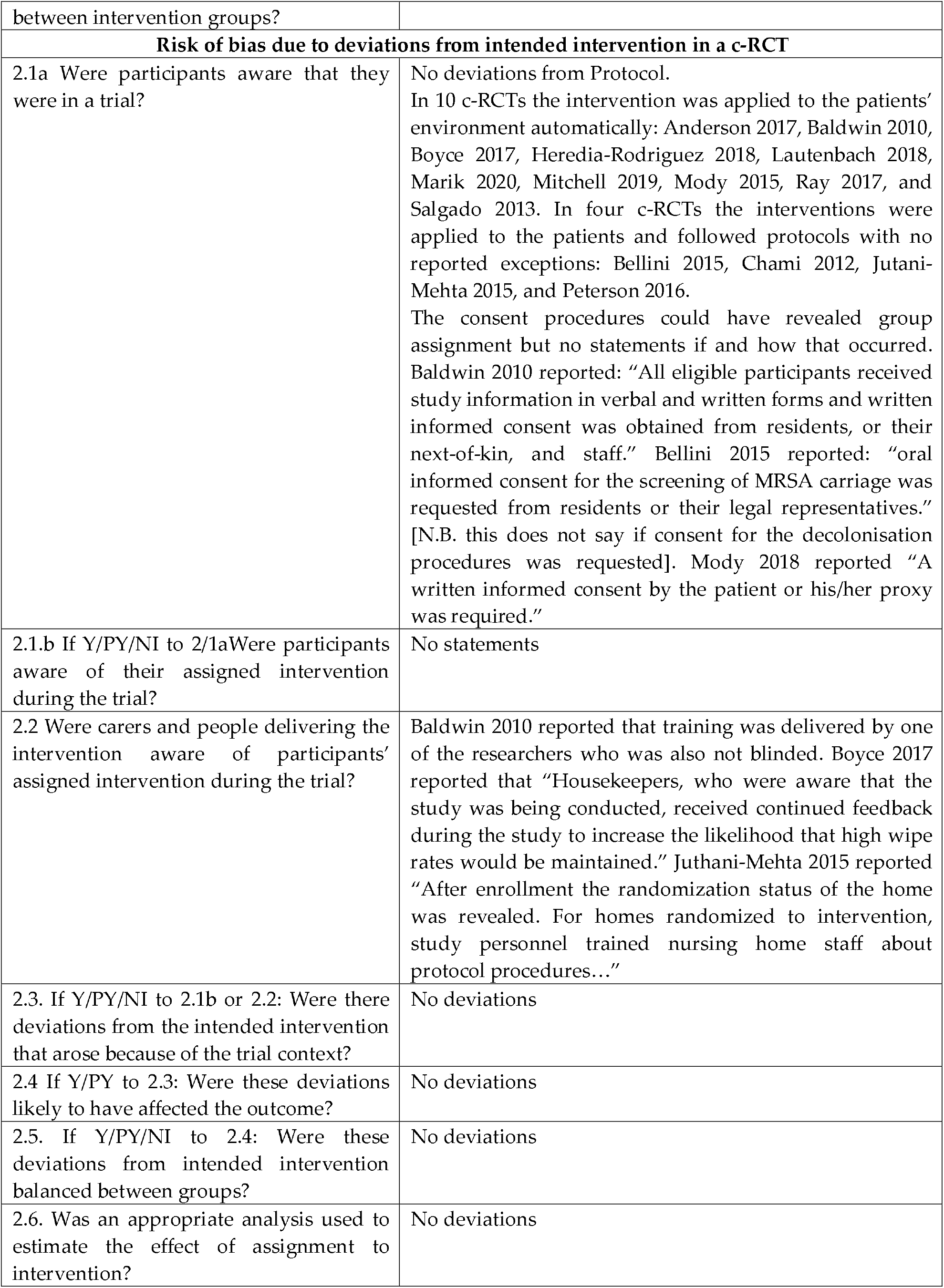
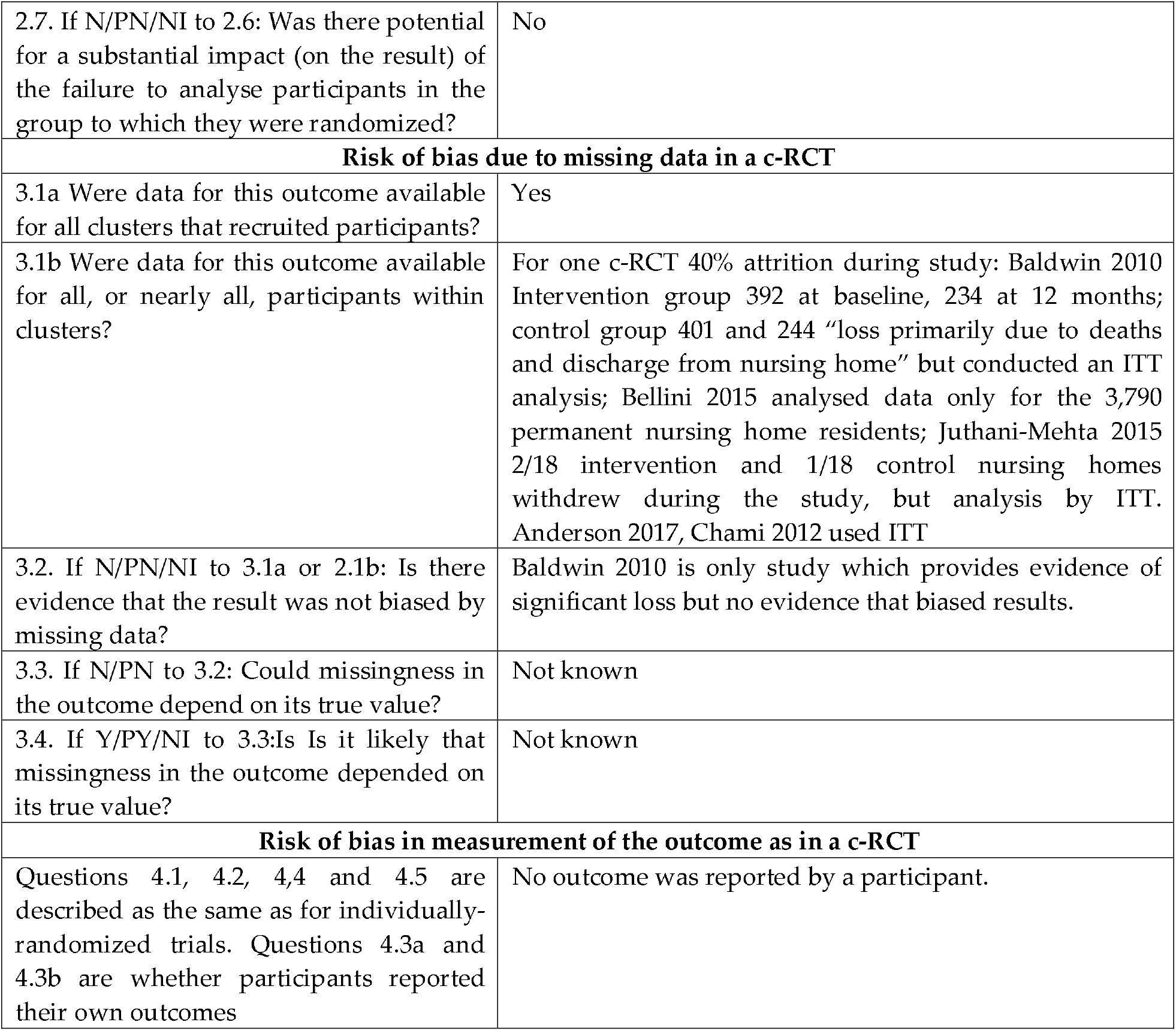

### Appendix C. GRADE assessment

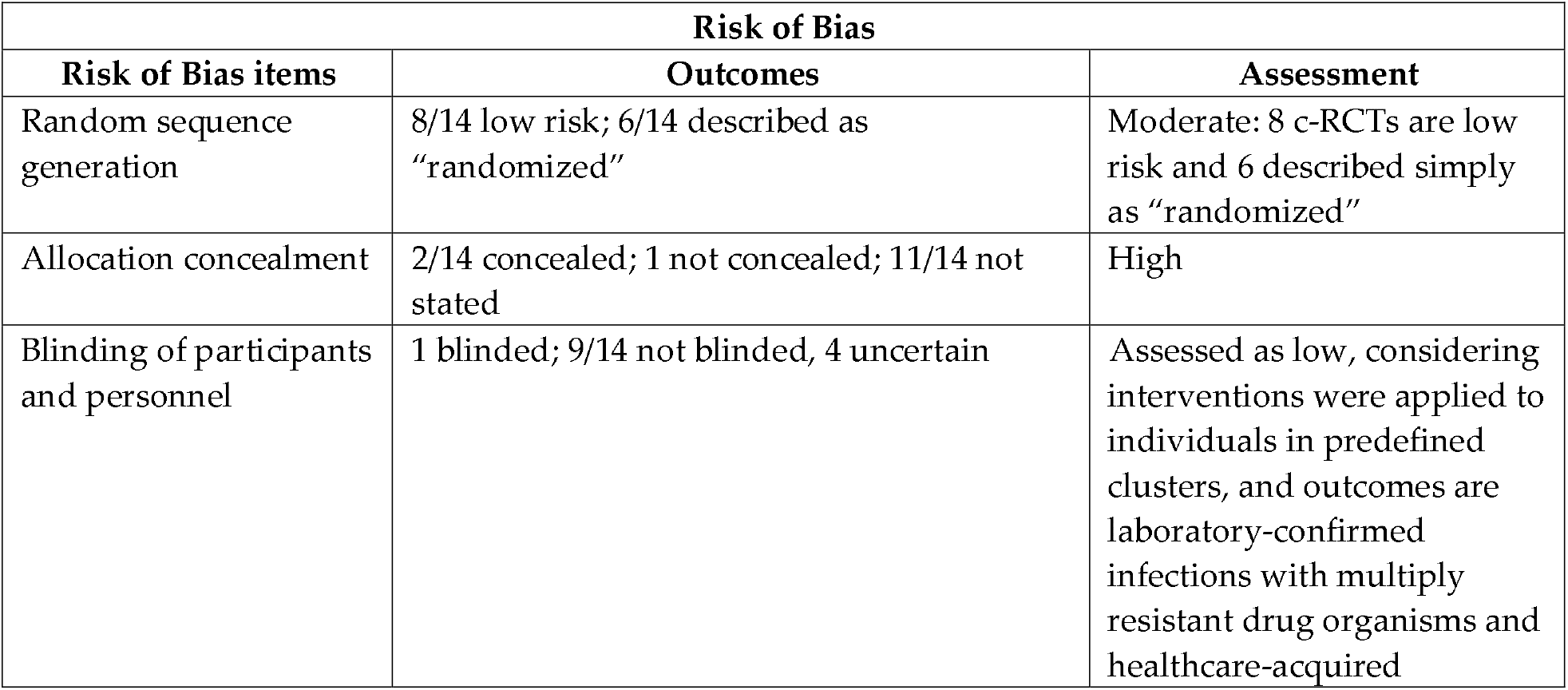
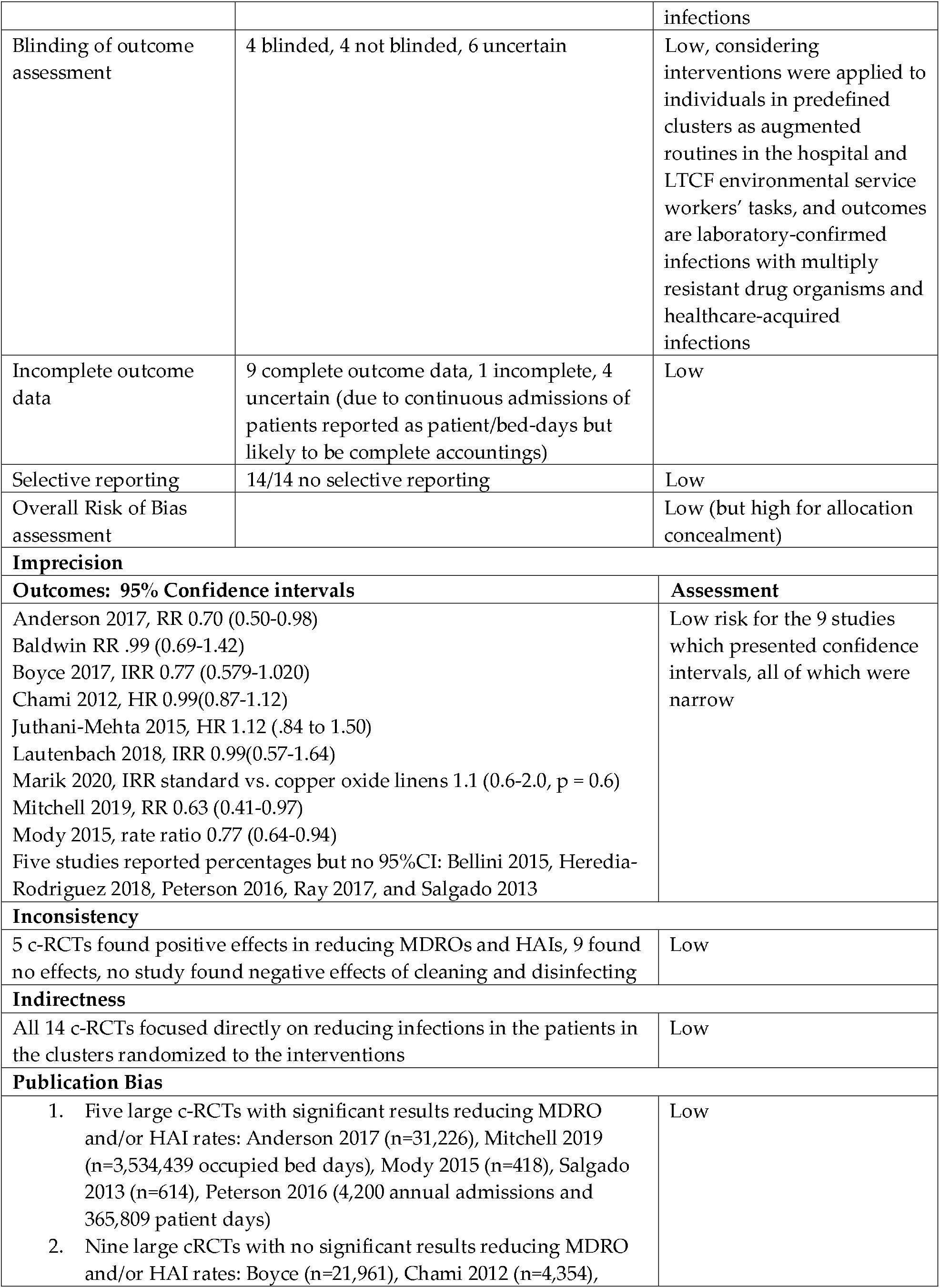
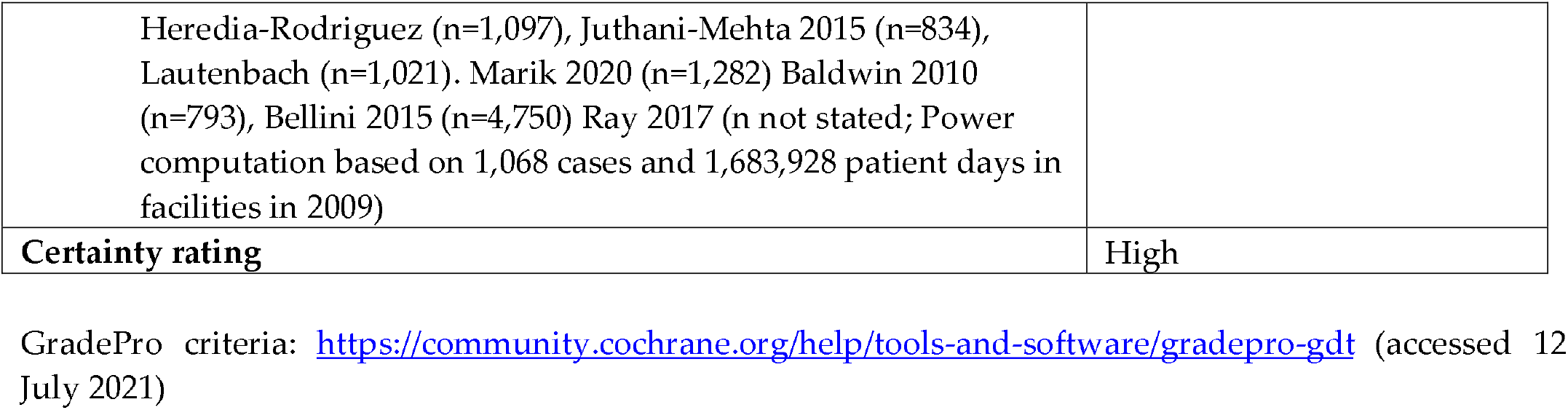

| Risk of Bias |  |  |
| --- | --- | --- |
| Risk of Bias items | Outcomes | Assessment |
| Random sequence generation | 8/14 low risk; 6/14 described as "randomized" | Moderate: 8 c-RCTs are low risk and 6 described simply as "randomized" |
| Allocation concealment | 2/14 concealed; 1 not concealed; 11/14 not stated | High |
| Blinding of participants and personnel | 1 blinded; 9/14 not blinded, 4 uncertain | Assessed as low, considering interventions were applied to individuals in predefined clusters, and outcomes are laboratory-confirmed infections with multiply resistant drug organisms and healthcare-acquired |
|  |  | infections |
| Blinding of outcome assessment | 4 blinded, 4 not blinded, 6 uncertain | Low, considering interventions were applied to individuals in predefined clusters as augmented routines in the hospital and LTCF environmental service workers' tasks, and outcomes are laboratory-confirmed infections with multiply resistant drug organisms and healthcare-acquired infections |
| Incomplete outcome data | 9 complete outcome data, 1 incomplete, 4 uncertain (due to continuous admissions of patients reported as patient/bed-days but likely to be complete accountings) | Low |
| Selective reporting | 14/14 no selective reporting | Low |
| Overall Risk of Bias assessment |  | Low (but high for allocation concealment) |
| **Imprecision** |  |  |
| **Outcomes: 95% Confidence intervals** |  | **Assessment** |
| Anderson 2017, RR 0.70 (0.50-0.98)<br>Baldwin RR .99 (0.69-1.42)<br>Boyce 2017, IRR 0.77 (0.579-1.020)<br>Chami 2012, HR 0.99(0.87-1.12)<br>Juthani-Mehta 2015, HR 1.12 (.84 to 1.50)<br>Lautenbach 2018, IRR 0.99(0.57-1.64)<br>Marik 2020, IRR standard vs. copper oxide linens 1.1 (0.6-2.0, p = 0.6)<br>Mitchell 2019, RR 0.63 (0.41-0.97)<br>Mody 2015, rate ratio 0.77 (0.64-0.94)<br>Five studies reported percentages but no 95%CI: Bellini 2015, Heredia-Rodriguez 2018, Peterson 2016, Ray 2017, and Salgado 2013 |  | Low risk for the 9 studies which presented confidence intervals, all of which were narrow |
| **Inconsistency** |  |  |
| 5 c-RCTs found positive effects in reducing MDROs and HAIs, 9 found no effects, no study found negative effects of cleaning and disinfecting |  | Low |
| **Indirectness** |  |  |
| All 14 c-RCTs focused directly on reducing infections in the patients in the clusters randomized to the interventions |  | Low |
| **Publication Bias** |  |  |
| Five large c-RCTs with significant results reducing MDRO and/or HAI rates: Anderson 2017 (n=31,226), Mitchell 2019 (n=3,534,439 occupied bed days), Mody 2015 (n=418), Salgado 2013 (n=614), Peterson 2016 (4,200 annual admissions and 365,809 patient days)<br>Nine large cRCTs with no significant results reducing MDRO and/or HAI rates: Boyce (n=21,961), Chami 2012 (n=4,354), |  | Low |

|  |  |
| --- | --- |
| Heredia-Rodriguez (n=1,097), Juthani-Mehta 2015 (n=834), Lautenbach (n=1,021). Marik 2020 (n=1,282) Baldwin 2010 (n=793), Bellini 2015 (n=4,750) Ray 2017 (n not stated; Power computation based on 1,068 cases and 1,683,928 patient days in facilities in 2009) |  |
| **Certainty rating** | High |

GradePro criteria: https://community.cochrane.org/help/tools-and-software/grade-pro-gdt (accessed 12 July 2021)

