## Supplementary material for "Cleaning and disinfecting surfaces in hospitals and long-term care facilities for reducing hospital and facility-acquired bacterial and viral infections: A systematic review": PRISMA Statement

| **Section and Topic** | **Item #** | **Checklist item** | **Location where item is reported** |
| --- | --- | --- | --- |
| **TITLE** | | |  |
| Title | 1 | Cleaning and disinfecting surfaces in hospitals and long-term care facilities and reducing bacterial and viral infections in patients: A systematic review. | 1 |
| **ABSTRACT** | | |  |
| Abstract | 2 | **Background:** A key concern of health care professionals and institutions is preventing patients acquiring infections, particularly multiply drug-resistant organisms (MDROs): methicillin-resistant *Staphylococcus aureus*, vancomycin-resistant enterococcus, *C. difficile*, *Acinetobacter* species, and extended spectrum beta-lactamase producing organisms, or Healthcare-Associated Infections (HAIs).  **Methods:** CINAHL, Cochrane CENTRAL Register of Controlled Trials, EMBASE, Medline, and Scopus were searched from inception to June 28, 2021. Searches combined terms for: (a) hospitals and acute/long term care facilities, (b) disinfectants (e.g: antisepsis, bleach, cleaning, copper plating of surfaces and copper impregnation of textiles, disinfection, decontamination, hydrogen peroxide, quaternary ammonium compounds, ultraviolet light), and antimicrobial-treated surfaces; and (c) RCTs, using the Cochrane Collaboration’s highly sensitive search filter. No language or date limits. Study reference lists were searched. Searches were downloaded into Covidence™ to remove duplicates and two authors independently screened abstracts and full-text papers and extracted data, with disagreements resolved by consensus. Studies were included if they: (1) focused on patients in hospitals or LTCFs; (2) reported on interventions to use bleach, quaternary ammonium disinfectants, hydrogen peroxide, ultraviolet lights, or copper-plated surfaces/copper-treated fabrics or antiseptic coated surfaces to disinfect surfaces or reduce colonisation rates in patients; (3) reported patient infections with MDROs and/or HAIs; and (4) were RCTs or c-RCTs. The Cochrane Risk of Bias Tool (RoB) and the Risk of Bias tool version 2 for cluster-randomised controlled trials (RoB v 2) were used to assess the risk of bias of c-RCTs included in this review, with disagreements resolved by consensus.  **Results:** Fourteen c-RCTs were identified in hospitals and LTCFs. The interventions in ten focused on reducing patient infections with MDROs and/or HAIs and four found significant reductions in patient infection rates with strategies including bleach, quaternary ammonium detergents, ultraviolet lights and hydrogen peroxide vapour. Three focused on reducing MRSA rates and one had significant results and one c-RCT focused on reducing *C. difficile*, rates and had no significant results. Heterogeneity in populations, methods, outcome measures and data reporting precluded meta-analysis.  **Discussion:** Key steps in reducing bacterial and viral infections are vaccinating all staff and patients, encouraging visitor vaccination, screening all admissions and discharges between institutions, isolating appropriate individuals until cleared, improving disinfection techniques of the identified studies, increasing antibiotic stewardship, more education and pay for cleaning staff and career paths teaching colleagues infection control. |  |
| **INTRODUCTION** | | |  |
| Rationale | 3 | Hospitals and LCTFs are open institutions with many new admissions and discharges. Pathogens circulate between hospitals, LTCFs and the community. To prevent new infections entering hospitals and LTCFs, infecting residents and staff and then being transmitted to other hospitals or LTCFs or the community and being important causes of morbidity and mortality the key question is which are the most evidence-based techniques to prevent transmission between patients, staff, visitors and community members by cleaning and disinfecting surfaces? |  |
| Objectives | 4 | **Population:** Patients in hospitals and LCTFs. **Interventions:** c-RCTs and RCTs of interventions using bleach, quaternary ammonium disinfectants, liquid or vapourised hydrogen peroxide, ultraviolet lights, copper-plated surfaces or copper-treated fabrics, or disinfectant or antiseptic coated surfaces. **Comparison:** usual cleaning methods in the institution (or quaternary ammonium disinfectant if it is the usual method). **Outcomes:** rates of new multiply drug-resistant infections (MDROs) and Healthcare-Associated Infections (HAIs) in patients.  **Secondary outcomes:** Secondary outcomes were whether the c-RCTs: (1) reported trends in MDRO rates in their region, (2) followed or tested evidence-based disinfection guidelines, (3) performed genomic studies to track the transmission of pathogens between the community, hospitals and LTCFs, (4) reported antibiotic stewardship programmes, and (5) measured MDRO and HAI rates in environmental service workers (ESWs, also called cleaners) and other HCWs. |  |
| **METHODS** | | |  |
| Eligibility criteria | 5 | C-RCTs and RCTs of interventions to decolonise patients and disinfect surfaces from bacteria and viruses in hospitals and LCTFs. No limitations of date and language. Exclude non-randomised studies. |  |
| Information sources | 6 | CINAHL, Cochrane CENTRAL Register of Controlled Trials, EMBASE, Medline, and Scopus were searched from inception to June 28, 2021. No language or date limits. Study reference lists were searched. |  |
| Search strategy | 7 | Five multidisciplinary databases (CINAHL, Cochrane CENTRAL Register of Controlled Trials, EMBASE, Medline, and Scopus) were searched from inception to June 28, 2021. Searches combined terms from: (a) hospitals and acute/long term care facilities (e.g: assisted living, long term care facilities, nursing homes) (b) disinfectants (e.g: antisepsis, bleach, cleaning, copper plating of surfaces and copper impregnation of textiles, disinfection, decontamination, hydrogen peroxide, quaternary ammonium compounds, ultraviolet light and antimicrobial treatment of surfaces); and (c) randomized controlled trials using the Cochrane Collaboration’s highly sensitive search filter for RCTs. Terms were searched as both keywords and database-specific subject headings to identify relevant RCTs. Systematic reviews and meta-analyses were also sought. |  |
| Selection process | 8 | Inclusion criteria were RCT or c-RCT and exclusion criteria non-randomised studies. Two reviewers independently reviewed all titles and abstracts. |  |
| Data collection process | 9 | Data on study populations, interventions, outcomes and analysis methods were abstracted independently by two reviewers. |  |
| Data items | 10a | Interventions (bleach, disinfectant, ultraviolet light, liquid or vapourised hydrogen peroxide, copper-treated surfaces or fabrics, antimicrobial treated surfaces); cleaning protocols and methods; protocol adherence; hand hygiene adherence of cleaners; outcomes (patient bacterial and viral infections, bacterial and viral contamination of surfaces). |  |
|  | 10b | Methods of assessing patient infection by laboratory studies including cultures, tests such as RT-PCR, and imaging; surface contamination by bacteriological examination of swabs pre – and post-infection; and reporting on bacterial rates and colony-forming units (cfus)/surface area. No methods were used to replace missing data. |  |
| Study risk of bias assessment | 11 | Cochrane risk of bias (RoB) tool and RoB tool version 2 for c -RCTs applied independently by two reviewers. |  |
| Effect measures | 12 | Authors used a variety of outcome measures (healthcare-associated infections (HAIs), multiply drug resistant organisms (MDROs), cfus/surface area), and reported them as ORs, RRs, or IRRs with 95% confidence intervals. Studies were heterogenous in study populations, infections studied, cleaning methods, infection outcomes and statistical reporting of outcomes and thus there were insufficient data arms for meta-analyses. |  |
| Synthesis methods | 13a | Insufficient data for meta-analyses. |  |
|  | 13b | No replacement of missing data, no data conversions. |  |
|  | 13c | Heterogeneous studies, insufficient data for meta-analyses. |  |
|  | 13d | Studies were summarised according to study design, methods of detecting infections, decolonisation methods and outcomes for patients and colonisation of surfaces. |  |
|  | 13e | Heterogeneous studies, insufficient data for meta-analyses. |  |
|  | 13f | Heterogeneous studies, insufficient data for meta-analyses thus no sensitivity analyses |  |
| Reporting bias assessment | 14 | **Risk of Bias:** Using the Cochrane Risk of Bias Tool, of the 12 c-RCTs with clinical patient outcomes, six were at low risk of bias for randomisation (all used computer programmes) and six were unknown as they merely stated they were randomised. Two were at low risk for allocation concealment, nine at unknown risk and one at high risk. The cleaning and disinfection interventions were applied by the environmental service workers employed by the hospitals and LTCFs to room surfaces and not to patients. The thoroughness of disinfection was assessed by environmental service supervisors (ESS) employed by the hospitals and LTCFs and also by the research staff and feedback was provided by the ESSs to the environmental service workers. Thus allocation concealment, though usually regarded as a key risk of bias concern, may be of less concern because of the use of hospital personnel in augmented disinfection routines. None were at low risk for blinding of participants and personnel risk, four were unknown and eight were at high risk; four were at low risk for blinding of outcome assessors, four were unknown and four were at high risk. However, for both blinding assessments, considering interventions were applied to all surfaces in predefined clusters using hospital and LTCF patient registries, and outcomes are laboratory-confirmed infections with MDROs or HAIs the risk of bias due to blinding is revised to low. Eight were considered at low risk for attrition and four were at unknown risk. Although several very large studies had a constant throughput of patients during the study period, the tracking of patients by patient bed occupancy which used the hospital admission and discharge system is likely to find all participants. All 12 studies were at low risk of selective reporting (Figures 1 and 2). The overall assessment is low risk but high for allocation concealment.  For the four c-RCTs which reported patient colonization rates Boyce [14] and Ray [30] reported patient outcomes (reported above). For randomization three were at low risk and one unknown, for allocation concealment four were at unknown risk, for blinding of participants and personnel one was at low risk and three at high risk, for blinding of outcome assessors one was at low risk, two unknown and one at high risk, for incomplete data one was at low risk two unknown and one at high risk, and for selective reporting all four were at low risk. The overall assessment was low risk but risk was high for allocation concealment. The same comments that the interventions were provided to predetermined clusters of wards apply but four studies required ongoing patient cooperation for personal decolonisation measures [15,16, 27,28].  **Imprecision:** Low (Low risk for the 9 studies which presented confidence intervals, all of which were narrow)  **Inconsistency:** Low (5 c-RCTs found positive effects in reducing MDROs and HAIs, 9 found no effects, no study found negative effects of cleaning and disinfecting)  **Indirectness:** Low  **Publication Bias:** Low |  |
| Certainty assessment | 15 | High |  |
| **RESULTS** | | |  |
| Study selection | 16a | Please see Item 7 above and Figure 1. |  |
|  | 16b | No studies that met the inclusion criteria were subsequently excluded. |  |
| Study characteristics | 17 | Of the 14 c-RCTs that reported patient infections, 10 assessed the effects of interventions on multiple MDROs or HAIs; Four studies assessed the effect of interventions on single MDROs (3 studies of methicillin-resistant *Staphylococcus aureus* (MRSA) and one of *C. difficile*). |  |
| Risk of bias in studies | 18 | Of the 14 c-RCTs with clinical patient outcomes eight were at low risk of bias for randomisation (all used computer programmes) and six were unknown as they merely stated they were randomised. Two were at low risk for allocation concealment, 11 at unknown risk and one at high risk. One was at low risk for blinding of participants and personnel risk, four unknown and nine were at high risk; four at low risk for blinding of outcome assessors, six unknown and four at high risk. Nine were at low risk for attrition, four unknown and one at high risk (several large studies reported patient bed occupation days and had a constant throughput of patients during the study period). None were at risk of selective reporting.  **Secondary outcomes:** (1) **MRDO trends:** only one c-RCT reported trends in MDRO rates in their region (Bellini 2015 for eastern Switzerland], (2) **Guidelines:** four c-RCTs referenced guidelines. Mitchell (2019) assessed their pragmatic research design against the PRagmatic-Explanatory Continuum Indicator tool and based the high touch points sampled on the CDC Environmental Cleaning Checklist. Chami (2012) presented during HCW training a “Delphi web survey of guidelines.” Mody (2015) reported all Medicare-certified and Medicaid-certified NHs have an infection control program, and Salgado (2013) referenced the CDC Guideline for Disinfection and Sterilization in Healthcare Facilities 2008 but the study intervention was copper surfaces, a new intervention, (3) **Genomic studies:** one c-RCT reported a genomic study that in each of the three LTCFs the MRSA clone (pulsotype) USA 100 predominated, (4) **Antibiotic stewardship:** only one study (Mitchel 2019) reported that all hospitals had an antibiotic stewardship programme throughout the study period but did not report its results, and (5) **HCW MDRO rates:** only one c-RCT (Baldwin 2010) reported MDRO rates in HCWs. |  |
| Results of individual studies | 19 | Of the 10 studies with interventions planned to reduce multiple MDROs four had significant reductions in patient infection rates. Anderson (2017) found the lowest rates of MDROs for the new admissions were after using ultraviolet light (UV) + quaternary ammonia detergent (QUAD) with 33.9 cases/10,000 exposure days, then after cleaning with bleach 41.6 cases/10,000 exposure days, then after UV + bleach 45.6 cases/10,000 exposure days and then for the reference study arm (QUAD) 51.3 cases/10,000 patient days. For *C. difficile* for bleach there were 30.4 cases/10,000 exposure days and for UV + bleach there were 31.6 cases/10,000 exposure days.  Mitchell (2019) found a significant reduction in VRE from 0.35/1,000 occupied bed days to 0.22 but no significant decrease in MRSA or *C. difficile.*  Salgado (2013) found for the combined outcome of HAI or colonisation with MRSA or VRE the rate was significantly lower in copper-plated rooms (0.071 vs. 0.123, p = 0.02) than rooms without copper plating of surfaces.  Mody (2015) found significant reductions in the intervention group for three outcomes: MRSA (RR 0.78; 95%CI 0.64 to 0.96), first catheter-associated urinary tract infections (RR 0.54, 95%CI 0.30 to 0.97) and all catheter-associated infections (RR 0.69, 95%CI 0.49 to 0.99) but no reductions in four outcomes: new VRE or resistant gram-negative bacilli infections, or new feeding tube–associated pneumonias or skin and soft-tissue infections.  Peterson (2016) found the baseline MRSA colonisation rate was 16.84% and after one year 11.61% in the intervention units (p = 0.028) and after one year 17.85% in the control units (p = 0.61). |  |
| Results of syntheses | 20a | Heterogeneous studies, insufficient data for meta-analyses. |  |
|  | 20b | Heterogeneous studies, insufficient data for meta-analyses. |  |
|  | 20c | Heterogeneous studies, insufficient data for meta-analyses. |  |
|  | 20d | No sensitivity analyses because no meta-analysis. |  |
| Reporting biases | 21 | For the studies which did not report the total number of patients and instead reported the throughput of patients the outcomes were reported per 1000 occupied bed days and risk of bias due to missing results was not assessed. |  |
| Certainty of evidence | 22 | High certainty. Five of 14 studies found significant reductions in patient infections. Four of these studies focused on reductions in MDROs and/or HAIs and one on MRSA. Nine studies had no significant reductions, and no study had an increase in infections. |  |
| **DISCUSSION** | | |  |
| Discussion | 23a | Key steps in reducing bacterial and viral infections are identifying patterns of transmission of MDROs and HAIs between hospitals, LCTFs and the communities in a region, increasing antibiotic stewardship and supervision of antimicrobial use in institutions and the community, screening all admissions and discharges between institutions, isolating appropriate individuals in isolation rooms until cleared, improving cleaning and disinfection techniques by detailed studies of environmental service workers’ (ESWs) training, supervision, and measurement of their cleaning and disinfecting skills, more education and pay for ESW staff and career paths teaching ESWs and other HCWs infection control. |  |
|  | 23b | Ten c-RCTs studied several MDROs and HAIs, three only MRSA and one only *C. difficile*. Studies varied in the cleaning methods and substances they used, the type and number of surfaces cleaned, whether they checked cleaning effectiveness by invisibly marking surfaces before cleaning, the proportion of cleaned surfaces sampled after cleaning, and the outcome statistics reported. Three studies (Boyce 2017, Mitchell 2919, and Peterson 2016) did not report patient numbers but patient occupied bed days. |  |
|  | 23c | The heterogeneity of methods, outcome measures and statistical analyses precluded meta-analysis because they resulted in insufficient similar study arms for meta-analysis. |  |
|  | 23d | Key steps are identifying symptomatic and asymptomatic individuals with MDROs and HAIs in hospitals, LTCFs and the community, and reducing their risk of transmission to other individuals by treatment and if necessary temporary isolation until they are cleared. The transmission of MDROs and HAIs needs to be identified by cultures, contact tracing and genomic analysis for the entire group of facilities which exchange and admit patients. Increased research is needed into improving the effectiveness of cleaning and disinfecting methods for all surfaces and all medical equipment, the synergy between preventing respiratory, person-to-person and person-to-surfaces transmission, and the training and effectiveness of cleaners. |  |
| **OTHER INFORMATION** | | |  |
| Registration and protocol | 24a | Prospero registration number is CRD42021249823. |  |
|  | 24b | On the Prospero website |  |
|  | 24c | The Prospero title included meta-analysis, but after completing the systematic review there were insufficient studies to permit meta-analysis. |  |
| Support | 25 | No financial support for the review. |  |
| Competing interests | 26 | No competing interests of review authors. |  |
| Availability of data, code and other materials | 27 | Data used for all analyses available from authors. |  |

*From:* Page MJ, McKenzie JE, Bossuyt PM, Boutron I, Hoffmann TC, Mulrow CD, et al. The PRISMA 2020 statement: an updated guideline for reporting systematic reviews. BMJ 2021;372:n71. doi: 10.1136/bmj.n71

For more information, visit: <http://www.prisma-statement.org/>
